# Leveraging cancer mutation data to predict the pathogenicity of germline missense variants

**DOI:** 10.1101/2024.03.11.24304106

**Authors:** Bushra Haque, David Cheerie, Amy Pan, Meredith Curtis, Thomas Nalpathamkalam, Jimmy Nguyen, Celine Salhab, Bhooma Thiruvahindrapura, Jade Zhang, Madeline Couse, Taila Hartley, Michelle M. Morrow, E Magda Price, Susan Walker, David Malkin, Frederick P. Roth, Gregory Costain

## Abstract

Innovative and easy-to-implement strategies are needed to improve the pathogenicity assessment of rare germline missense variants. Somatic cancer driver mutations identified through large-scale tumor sequencing studies often impact genes that are also associated with rare Mendelian disorders. The use of cancer mutation data to aid in the interpretation of germline missense variants, regardless of whether the gene is associated with a hereditary cancer predisposition syndrome or a non-cancer-related developmental disorder, has not been systematically assessed. We extracted putative cancer driver missense mutations from the Cancer Hotspots database and annotated them as germline variants, including presence/absence and classification in ClinVar. We trained two supervised learning models (logistic regression and random forest) to predict variant classifications of germline missense variants in ClinVar using Cancer Hotspot data (training dataset). The performance of each model was evaluated with an independent test dataset generated in part from searching public and private genome-wide sequencing datasets from ∼1.5 million individuals. Of the 2,447 cancer mutations, 691 corresponding germline variants had been previously classified in ClinVar: 426 (61.6%) as likely pathogenic/pathogenic, 261 (37.8%) as uncertain significance, and 4 (0.6%) as likely benign/benign. The odds ratio for a likely pathogenic/pathogenic classification in ClinVar was 28.3 (95% confidence interval: 24.2-33.1, p < 0.001), compared with all other germline missense variants in the same 216 genes. Both supervised learning models showed high correlation with pathogenicity assessments in the training dataset. There was high area under precision-recall curve values of 0.847 and 0.829 for logistic regression and random forest models, respectively, when applied to the test dataset. With the use of cancer and germline datasets and supervised learning techniques, our study shows that cancer mutation data can be leveraged to improve the interpretation of germline missense variation potentially causing rare Mendelian disorders.

**AUTHOR SUMMARY:** Our study introduces an approach to improve the interpretation of rare genetic variation, specifically missense variants that can alter proteins and cause disease. We found that published evidence from somatic cancer sequencing studies may be relevant to understanding the impact of the same variant in the context of rare inherited (Mendelian) disorders. By using widely available datasets, we noted that many cancer driver mutations have also been observed as rare germline variants associated with inherited disorders. This intersection led us to employ machine learning techniques to assess how cancer mutation data can predict the pathogenicity of germline variants. We trained machine learning models and tested them on a separate dataset curated by searching public and private genome-wide sequencing data from over a million participants. Our models were able to successfully identify pathogenic genetic changes, demonstrating strong performance in predicting disease-causing variants. This study highlights that cancer mutation data can enhance the interpretation of rare missense variants, aiding in the diagnosis and understanding of rare diseases. Integrating this approach into current genetic classification frameworks could be beneficial, and opens new avenues for leveraging existing cancer research to benefit broader genetic research and diagnostics for rare genetic conditions.

## BACKGROUND

Genome-wide sequencing (GWS; including exome and genome sequencing) allows for comprehensive detection of coding sequence variants associated with a wide range of diseases, spanning from rare Mendelian disorders to common cancers.^1–3^ Our ability to filter and prioritize variants associated with disease lags behind our ability to detect variation.^2^ Rare missense variants are collectively common in every human genome,^3,4^ and interpreting the clinical impact of these variants is especially challenging. The American College of Medical Genetics and Genomics (ACMG) and the Association for Molecular Pathology (AMP) developed a widely used system for assessing variants by scoring lines of evidence supporting variant pathogenicity or benign-ness.^4^ Even after a decade of implementing and refining the ACMG/AMP classification system, variants of uncertain significance (VUS) account for the vast majority of missense variant entries in databases like ClinVar.^5,6^ Despite commendable efforts to generate functional data through multiplexed assays of variant effects (MAVEs) and other variant-to-function maps, missense variant classification in clinical practice continues to often rely on *in silico* evidence and heuristics like rarity and inheritance.^7,8^ New scalable and easy-to-implement strategies that produce evidence complementary to (and not derivative of) existing *in silico* methods are needed to improve the pathogenicity assessment of rare germline missense variants.

Using available but underused genomic databases to identify additional evidence for pathogenicity could aid in classifying rare missense variants.^8–10^ Oncogenic mutations (also known as cancer driver mutations) are genetic alterations that contribute to cancer initiation and progression.^11^ Tumour sequencing initiatives like The Cancer Genome Atlas (TCGA) and International Cancer Genome Consortium (ICGC) have accelerated the identification of oncogenic mutations.^3,12^ Germline dysregulation of some proto-oncogenes and tumour suppressor genes (TSGs) causes Mendelian disorders (“oncoprotein duality”) (Figure 1A).^7,11,13,14^ For instance, the somatic *HRAS*^Q61K^ missense mutation implicated in various types of cancers causes Costello syndrome (MIM #218040), a developmental disorder, when it occurs as a germline variant (Figure 1B).^15,16^ These Mendelian disorders may or may not include cancer as a major phenotypic feature.^5,17–21^ Walsh and colleagues previously explored the use of cancer mutational hotspots data for interpreting germline variants in genes causing cancer predisposition syndromes.^13^ However, when and to what extent cancer driver mutations are pathogenic in germline contexts, for rare Mendelian disorders in general, remains unknown.

**Figure 1.**
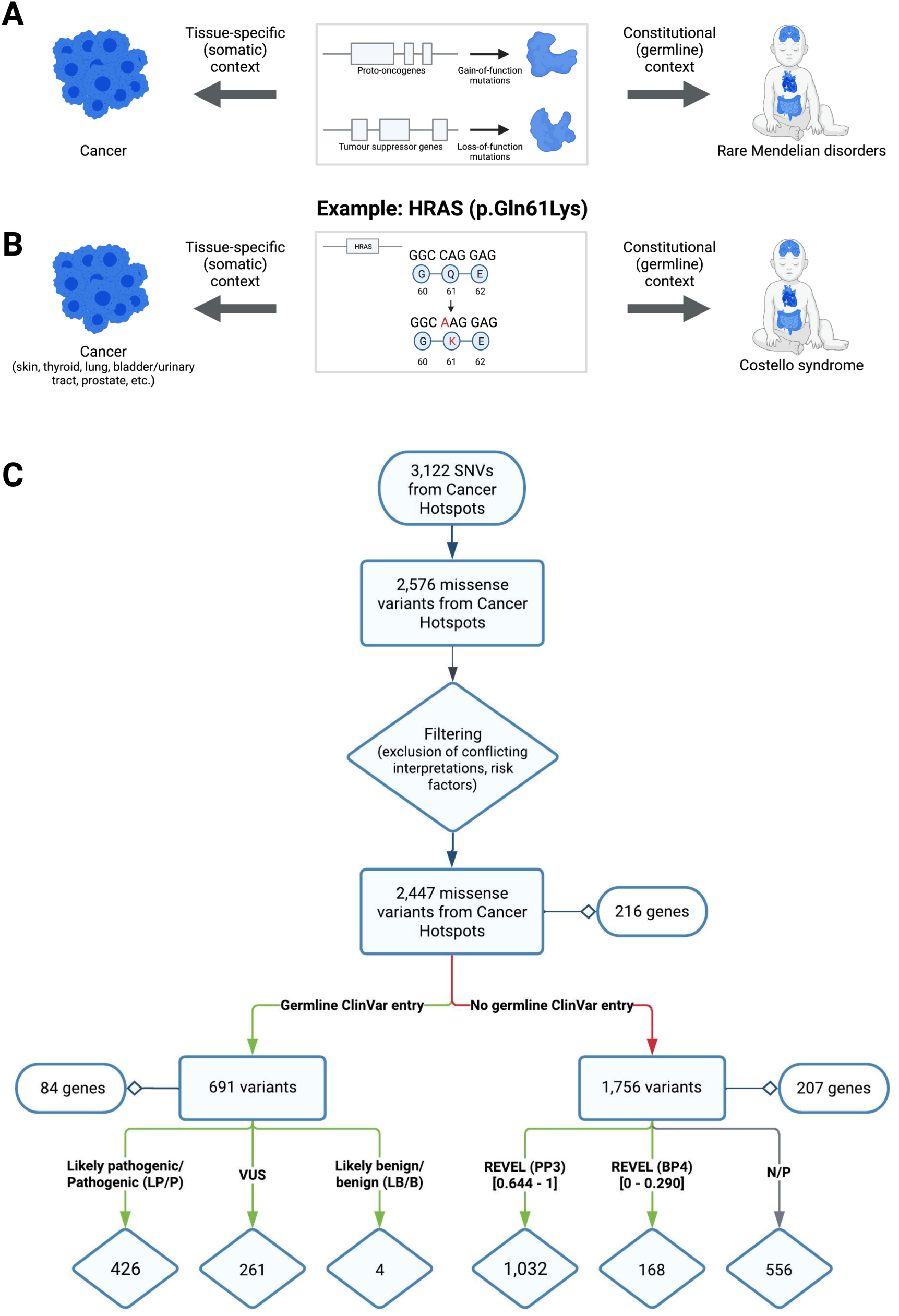
Germline variant and somatic cancer mutation overlap. (A) The presence of either gain-of-function or loss-of-function mutations in cancer driver genes can lead to cancer (left) or rare Mendelian disorders (right) in different contexts. Most cancers result from somatic mutations that accumulate in a tissue-specific manner, whereas germline mutations are present in all cells of the body and cause a type of rare Mendelian disorder (e.g., neurodevelopmental disorder). (B) The *HRAS*^Q61K^ mutation is an example of a known cancer mutation that drives different types of cancers that also causes Costello syndrome, a developmental disorder, when observed as a germline variant. (C) Workflow for extracting cancer mutations from Cancer Hotspots. Recurrent cancer mutations were filtered to 2,447 missense mutations. See main text for details. REVEL scores thresholds correspond to supporting evidence for pathogenicity (PP3) and for benign-ness (BP4). Created with Lucidchart.

This study investigates the concept of oncoprotein variant duality, and specifically the degree to which germline variant classification could be informed by observations that the equivalent tumour mutation drives cancer. The underlying logic of our approach is that cancer driver mutations have functional consequences at the protein level, and those functional consequences are expected to be present regardless of whether the variant is observed in a somatic/mosaic/tissue-specific or constitutional/germline context. Through comparative analysis of Cancer Hotspots^22,23^ (cancer mutations) and ClinVar^24^ (restricting to germline variants), we developed and tested supervised learning models for predicting germline missense variant pathogenicity using cancer mutation data.

## RESULTS

### Association between cancer mutations from Cancer Hotspots and LP/P classification as germline variants

Putative driver mutations from Cancer Hotspots were extracted, annotated, and filtered to obtain a list of 2,447 missense mutations (“CH mutations”) distributed across 216 genes (Figure 1C). Of these 216 genes, 41% are proto-oncogenes, 36% are tumour suppressor genes, and 15% can have either role, as determined by the Cancer Gene Census (Supplemental Figure 2A).^25^ We presumed that cancer driver missense mutations in proto-oncogenes and tumour suppressor genes have gain of function and loss of function mechanisms, respectively. The Mendelian disease associations in the Online Mendelian Inheritance in Man (OMIM) database^26^ for these genes revealed that 20% are associated with hereditary cancer predisposition syndromes (Supplemental Table 1). Among the 216 genes, 154 had known modes of inheritance for cancer and an associated Mendelian disease reported in OMIM.^26^ Of these 154 genes, 107 (69%) had a Mendelian disease mechanism that was concordant with the cancer mechanism, 26 (17%) were discordant, and 21 (14%) were semi-concordant, meaning the gene could function as both a proto-oncogene and a tumor suppressor, or had Mendelian diseases with variants exhibiting both gain of function and loss of function mechanisms (Supplemental Table 2). Although Cancer Hotspots infers cancer driver status of a mutation from probabilistic arguments (statistical enrichment), we found that the functional impact was experimentally tested for 990 of these mutations with the majority (943/990, 95%) confirmed to result in gain or loss of protein function (Supplemental Methods; Supplemental Figure 3).

Overall, 691 missense mutations in 84 genes from Cancer Hotspots had been classified with respect to germline pathogenicity in ClinVar: 426 (61.6%) as LP/P, 261 (37.8%) as VUS, and 4 (0.6%) as LB/B (Figure 1C). The median number of variants observed for each gene was 2 (interquartile range = 4). As expected, all variants were rare (gnomAD allele frequency < 0.001) except for three out of four that were classified as LB/B. Of these 84 genes, 50% are proto-oncogenes, 37% are tumour suppressor genes, and 10% can have either role, as determined by the Cancer Gene Census (Supplemental Figure 2B). Germline variants overlapping with cancer (driver) mutations may provide insights into their mechanisms, such as loss of function in tumor suppressor genes or gain of function in proto-oncogenes and provide functional context for Mendelian diseases. The disease associations in OMIM for these genes also revealed that 38% were hereditary cancer predisposition syndromes (e.g., *VHL* associated with von Hippel-Lindau syndrome) and 62% were not known to include cancer as a predominant feature (e.g., *FGFR3* associated with Achondroplasia).^26^ In both groups, most associated conditions had autosomal dominant inheritance (88% and 77%, respectively). A significant difference was observed in the proportion of LP/P, VUS, and LB/B variants between these two gene groups (256 LP/P, 231 VUS, 1 LB/B versus 170 LP/P, 30 VUS, 3 LB/B, respectively), with an LP/P classification more likely for variants in genes not associated with hereditary cancer predisposition syndromes (p < 2.2e-16) (Supplemental Table 1).

The odds ratio for these 691 variants having a LP/P classification in ClinVar was 107.6 (95% confidence interval (CI): 40.1-288.4, p < 0.0001), when comparing only LP/P and LB/B classifications with all other germline missense variants with ClinVar entries in the 216 genes (n=5,474) (Supplemental Figure 1; Supplemental Table 3). Even if all VUS were considered as LB/B variants, the odds ratio was 28.3 (95% CI: 24.2-33.1, p < 0.001) compared with all other variants in ClinVar (n=50,655) (Supplemental Figure 1; Supplemental Table 3). In an even more extreme scenario of considering all VUS and CIP variants as LB/B, the odds ratio was 21.0 (95% CI: 18.2-24.2, p < 0.001) (n=53,593) (Supplemental Figure 1; Supplemental Table 3). If these variants were restricted to the 107 genes with Mendelian disease mechanism that was concordant with the cancer mechanism, 337 cancer mutations would overlap with germline missense variants in ClinVar (238 LP/P, 98 VUS, 1 LB/B). The odds ratio for an LP/P classification in ClinVar would increase to 46.2 (95% confidence interval: 36.4 - 58.6, p < 0.001), compared to all other germline missense variants in the same 107 genes. However, the odds ratio for LP/P classification for the “discordant” and “semi-concordant” mechanisms was still 12.5 (95% confidence interval: 9.9 - 15.7, p < 0.001). The positive likelihood ratio of 11.5 exceeded “moderate evidence” thresholds described previously (i.e., 4.33 and 5.79) (Supplemental Table 4).^27,28^ The potential impact of an additional moderate evidence criterion for pathogenicity applied to the 261 CH mutations that overlap with germline VUS in ClinVar is shown in Supplemental Figure 4, revealing 66 (27%) of the VUS could be hypothetically upgraded to LP.

For the remaining CH mutations that did not overlap with germline variants in ClinVar (n = 1,756), we explored the degree to which *in silico* scores used for germline variant adjudication supported “pathogenicity”. We grouped these CH mutations by REVEL scores using the ClinGen-proposed PP3/BP4 score thresholds (Figure 1C).^28^ Over half (58.8%; 1,032) had REVEL scores indicating at least PP3-level evidence (i.e., evidence in favour of pathogenicity), while only 9.6% (168) had at least BP4-level evidence (Figure 1C; Supplemental Figure 5A). Findings were similar using AlphaMissense (Supplemental Figure 5B).^29^ For these CH mutations that are absent from ClinVar, the *in silico* score profiles resemble the ClinVar LP/P germline missense variants in the same genes more than the set of LB/B variants or VUS (Supplemental Figure 5).

Through collaborations with GEL, MSSNG, C4R, and GeneDx, we searched GWS datasets from approximately 1.5 million participants (probands and affected or unaffected family members) and identified additional instances of germline variants overlapping with CH mutations (Supplemental Table 5). Across the four datasets, we found 302 unique overlapping germline variants. Of these, 194 were already classified and present in ClinVar (140 LP/P, 1 LB/B, 53 VUS) and 108 were absent in ClinVar. Out of these 108 variants, 43 had been previously assessed and classified in accordance with ACMG/AMP variant interpretation guidelines by our collaborators. Among these variants, 30 were classified as LP/P, 12 as VUS, and 1 conflicting (LP and VUS by different groups). The classifications of the remaining 65 variants (79% found in probands) were uncertain due to limited phenotype information.

### Cancer Hotspots database includes most highly recurrent cancer mutations in COSMIC

We retrieved 231,377 somatic missense mutations by filtering the Cancer Census Genes data from COSMIC (Supplemental Figure 6). With the results of the tumour sample count analysis using overlapping CH mutations and ClinVar germline variants (Supplemental Methods, Supplemental Figure 7), we stringently filtered for COSMIC mutations that were observed in >25 tumour samples and absent from Cancer Hotspots, resulting in 125 missense mutations across 63 genes (Supplemental Figure 6). This approach, using Cancer Hotspots as a benchmark, aimed to identify recurrent (putative) driver mutations in COSMIC, a more heterogeneous database with both driver and passenger mutations. Of these genes, 31 are new additions to the list of genes from Cancer Hotspots and 11 are associated with rare Mendelian diseases as reported in OMIM.^26^ However, only 12 of these mutations overlapped with germline variants in ClinVar. Among them, 2 (16.7%) were LP/P, 8 (66.7%) VUS/CIP and 2 (16.7%) were LB/B (Supplemental Figure 6). Only 2 of these 12 overlapping variants were found in the “new” 31 cancer genes discovered through COSMIC. While we identified 125 additional missense mutations in COSMIC, only a small fraction of these overlapped with germline variants in ClinVar. Thus, despite being smaller and less frequently updated than COSMIC, Cancer Hotspots effectively captures most putative cancer driver missense mutations relevant to our research question.

### Robust predicted probabilities of pathogenicity generated by supervised learning models

We used the training datasets to develop two types of supervised learning models with the goal to accurately predict the pathogenicity of germline variants in our test dataset. The training dataset fit the LRM with a McFadden’s pseudo-R^2^ value of 0.50 (i.e., higher than the 0.20-0.40 range that indicates a good model fit^30^) and generated predicted probabilities of pathogenicity for all variants in the training dataset. The predicted probabilities were significantly higher for all germline LP/P variants compared with LB/B/VUS variants (U = 1655893, n_LB/B/VUS_ = 11,644, n_LP/P_ = 2,095, p < 0.0001) and for germline variants that are present in the Cancer Hotspots database compared with those that are absent (U = 32029, n_Absent_ = 13,316, n_Present_ = 423, p < 0.0001) (Figure 3AB). We trained a second supervised learning model, an RFM, since it is gene-independent and can be broadly applied to variants beyond the 66 gene categories in the LRM. The RFM achieved an out-of-bag (OOB) error estimate of 10.8% for predicting outcomes. The RFM generated probability scores of pathogenicity and, similar to the LRM, these were significantly higher for all germline LP/P variants compared with LB/B/VUS variants, as well as for germline variants that overlap with CH mutations compared to those without overlap (U = 6109589, n_LB/B/VUS_ = 11,644, n_LP/P_ = 2,095, p < 0.0001) (Figure 3CD). To gain a comprehensive understanding of the overall impact of each independent variable on the data, exploratory analyses were conducted on the ClinVar dataset (before filtering) (Supplemental Methods; Supplemental Figures 6-8). The analyses show variability in the number of variants across genes (Supplemental Figure 7), distinct tumour sample count thresholds between LP/P and LB/B/VUS variants (Supplemental Figure 8) and indicated that the model fit was not primarily driven by the conservation scores (Supplemental Figure 9).

**Figure 2.**
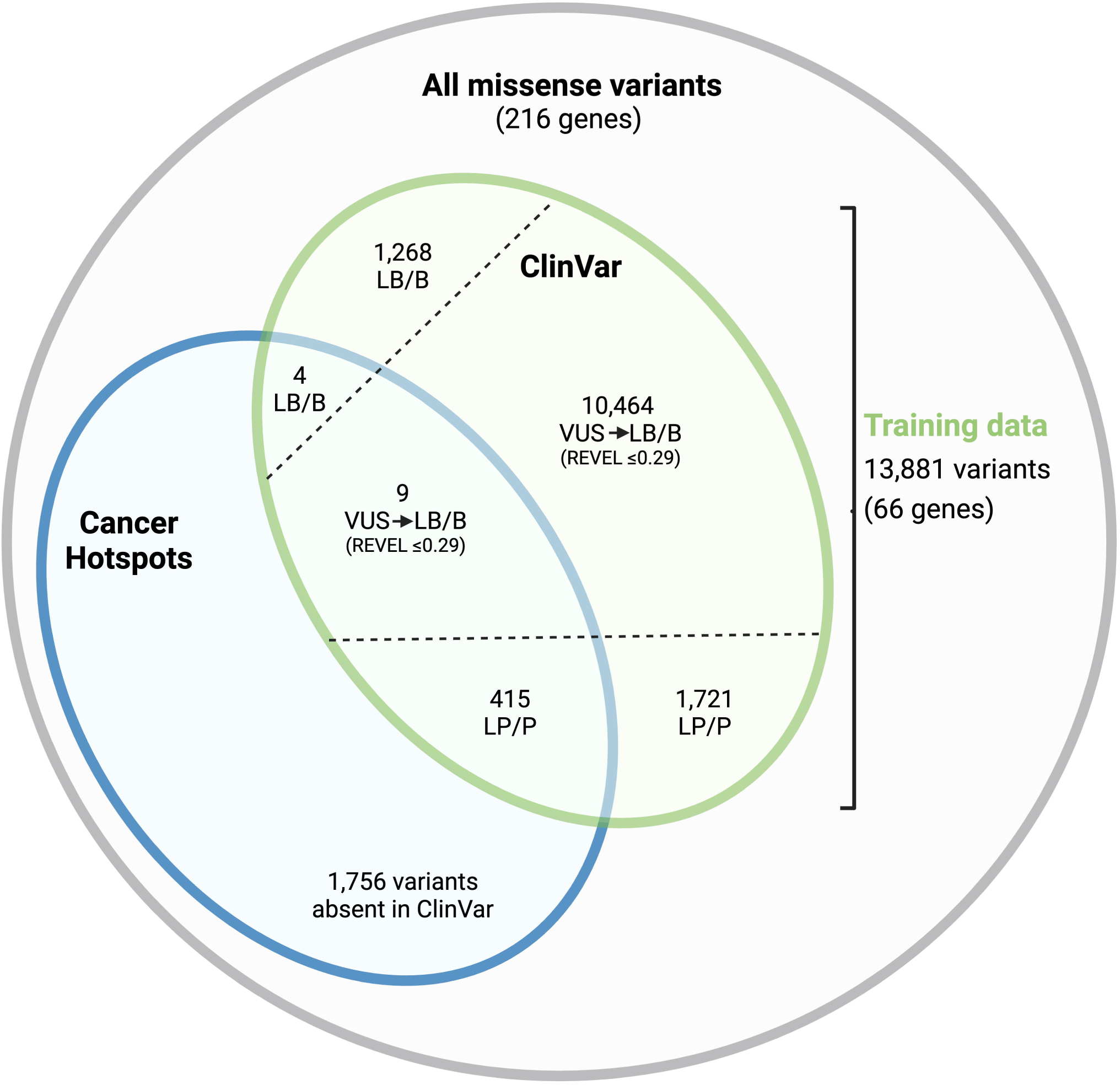
Training dataset for supervised learning models. The training dataset is comprised of 13,881 germline missense variants from ClinVar (green), including 691 overlapping with cancer mutations (blue). Different single nucleotide changes causing the same amino acid change were grouped together accounting for the difference in the overlap shown in Figure 1. Variants of uncertain significance (VUS) with REVEL scores ≤ 0.290 were included in the dataset and treated as likely benign/benign (LB/B) variants (see text for justification). LP/P, Likely pathogenic/Pathogenic. Created with BioRender.

**Figure 3.**
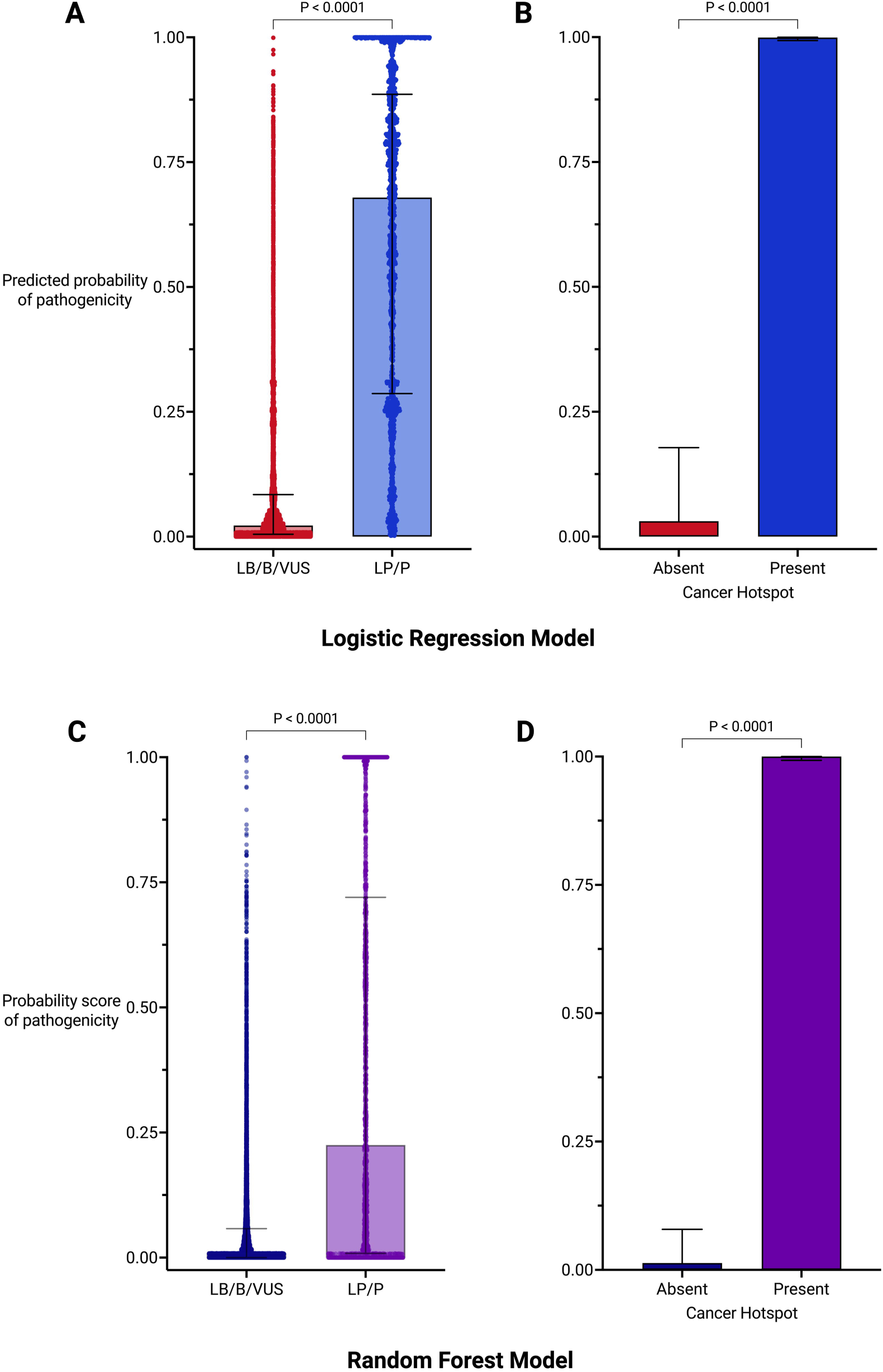
Fit of training dataset using supervised learning models. (A) Plot of predicted probabilities of pathogenicity for all likely benign/benign/variant of uncertain significance (LB/B/VUS) and likely pathogenic/pathogenic (LP/P) in the training dataset assigned by the logistic regression model. Mann-Whitney U test: U = 1655893, n_LB/B/VUS_ = 11,644, n_LP/P_ = 2,095. Comparison of predicted probabilities for germline variants with absence or presence of overlap with cancer mutations. Mann-Whitney U test: U = 32029, n_Absent_ = 13,316, n_Present_ = 423. Plot of probability scores of pathogenicity for LB/B/VUS and LP/P in the training dataset assigned by the random forest model. Mann-Whitney U test: U = 6109589, n_LB/B/VUS_ = 11,644, n_LP/P_ = 2,095. (D) Comparison of probability scores for germline variants with absence or presence of overlap with cancer mutations. Mann-Whitney U test: U =12913, n_Absent_ = 13,316, n_Present_ = 423. Created with GraphPad Prism.

### RFM outperformed LRM in correctly predicting pathogenicity of germline missense variants overlapping with cancer mutations

Using the test dataset (n = 332), distinct from training dataset variants, we calculated the area under precision-recall curve (AUPRC) values for the LRM and RFM as 0.847 and 0.829, respectively (Figure 4A). We also calculated the area under the receiver-operating characteristic curve (AUROC) as 0.821 for the LRM and 0.774 for the RFM (Supplemental Figure 10A). The higher AUROC for the LRM indicates better ability to discriminate between LP/P and LB/B/VUS variants compared to the RFM. Precision-recall curves guided the selection of optimal classification thresholds, with an emphasis on minimizing false positives while maximizing AUPRCs. The LRM had an optimal threshold of 0.74 (F1 score = 0.690) (Supplemental Figure 11A). The RFM had an optimal threshold of 0.39 (F1 score = 0.783) (Supplemental Figure 11B), with the higher F1 score compared with the LRM indicating superior performance in correctly predicting the pathogenicity of test dataset variants.

**Figure 4.**
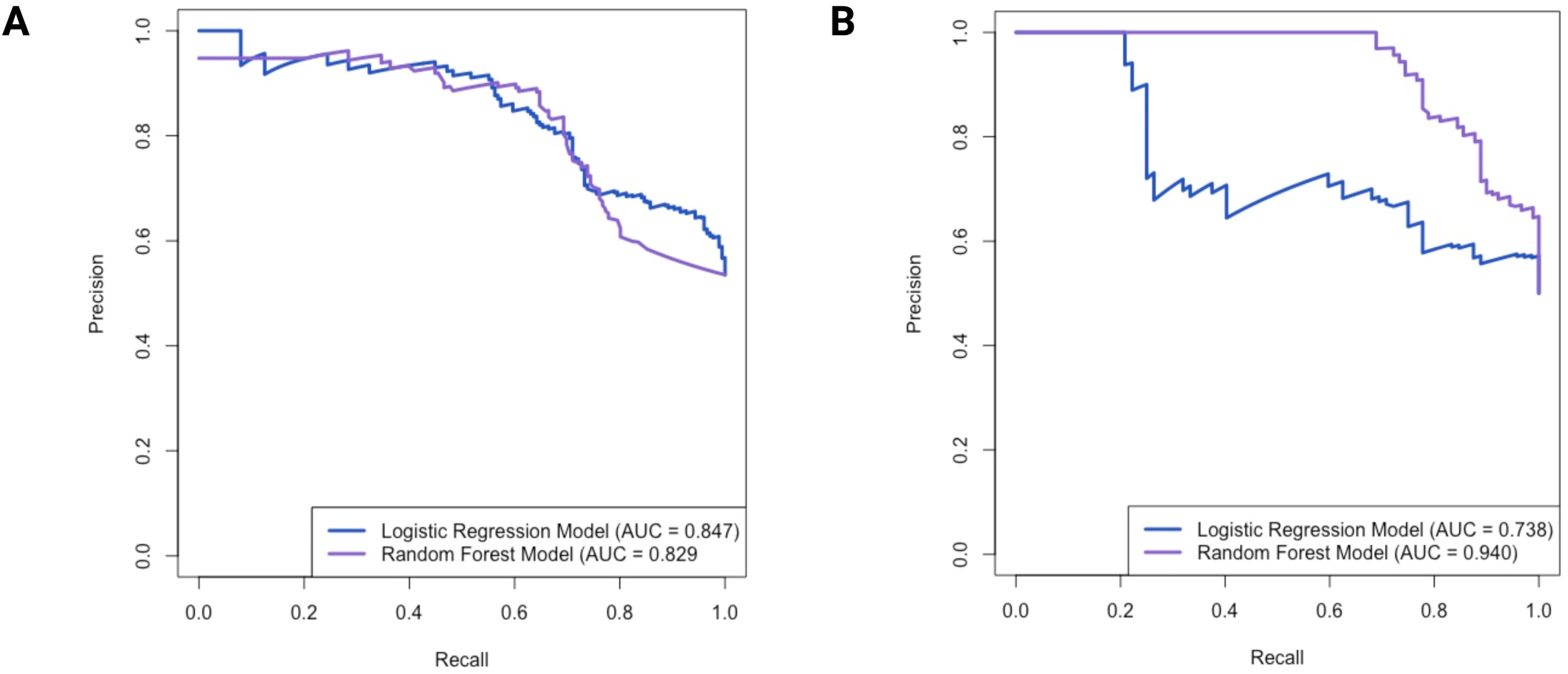
Evaluation of supervised learning models. Precision-recall curve comparing the performance of the logistic regression model (blue) and the random forest model (purple) using the (A) test dataset and (B) cross-validation set. The models’ performance was evaluated using k-fold cross-validation, with k=8 for logistic regression and k=10 for random forest. AUC, area under the curve.

We compared the performance of the LRM and RFM pathogenicity scores against the scores of other *in silico* prediction tools by plotting precision-recall curves and comparing the calculated AUPRCs (Supplemental Figure 12A). The LRM and RFM outperformed the first-generation tools^31^ SIFT and PolyPhen-2, which had AUPRCs of 0.821 and 0.827, respectively (Supplemental Figure 12B). Second- (REVEL, CADD, VARITY, VEST4) and third-generation (AlphaMissense, PrimateAI, MutPred2)^31^ tools demonstrated a stronger performance in classifying the test dataset variants, with AUPRCs ranging from 0.881 to 0.963 (Supplemental Figure 12CD). REVEL, VARITY, and AlphaMissense were the top-performing tools, respectively. Given the smaller size of the test dataset compared with the training dataset, cross-validation techniques were also used to confirm the LRM and RFM’s reliability in estimating performance (Figure 4B, Supplemental Figure 10B). The RFM consistently outperformed the LRM in terms of AUPRC, exhibiting a higher value than was observed with the test dataset alone (0.940 versus 0.738 AUC). Although the LRM had a higher AUROC (0.928) compared to the RFM (0.739), AUROC reflects overall discriminative ability across all thresholds, whereas AUPRC and F1 scores are more relevant for assessing performance in detecting positive cases. We used the RFM and the optimal threshold value of 0.39 to predict pathogenicity of the 65 variants with unknown classification identified through our collaborations with MSSNG, GEL, C4R, and GeneDx. Of these 65 variants, the RFM predicted 92% to be LP/P and 8% as LB/B. The average probability score of pathogenicity for the predicted LP/P variants was 0.93 and 80% were in probands.

## DISCUSSION

The increasing use of GWS in clinical practice has underscored the need for novel methods to interpret germline missense variation.^2,5,32^ We explored the generalizability of an understudied line of evidence that considers overlap with (presumed driver) cancer mutations. Using 2,447 cancer missense mutations from the Cancer Hotspots database, we identified significant enrichment for LP/P germline variants causing rare Mendelian disorders, regardless of cancer being or not being a major phenotype of the disorder. The results from our models support and extend these findings, by successfully predicting the pathogenicity of germline missense variants using supervised learning models trained with CH mutation data. Our findings indicate that statistically significant recurrent cancer mutation data can be leveraged to improve the interpretation of germline missense variation potentially causing rare Mendelian disorders.

Walsh and colleagues first proposed modifying the existing PM1 pathogenic evidence criterion to apply to germline variants in cancer predisposition genes that overlap with cancer mutations from Cancer Hotspots,^13^ provided the variant was not already in a germline hotspot.^4^ The results of our study support and extend this concept. A majority (62%) of genes considered in our study are not known to be associated with hereditary/germline cancer predisposition in a Mendelian disease context. We emphasize that this line of evidence is not codified in existing interpretation frameworks, including ACMG, ClinGen, and the Association for Clinical Genomic Science (ACGS), and is distinct from other criteria specific to missense variants, such as germline mutational hotspots (PM1) and instances where a previous pathogenic variant has been previously observed (PS1/PM5). This evidence may be most relevant in scenarios involving the interpretation of (rare) missense VUS. Cancer mutations may be embryonic lethal as germline variants;^11^ this biological constraint will limit the extent of overlap we observe between cancer mutations and germline variants.

The stand-alone probability scores of pathogenicity from our supervised learning models were not superior to other widely used *in silico* prediction tools in classifying germline missense variants. This was an expected result, since existing *in silico* tools were likely used *a priori* to inform classifications for these variants. Regardless, this comparison underscores our proposal that the LRM and RFM models would be used in addition to, rather than instead of, existing *in silico* tools for variant classification. Since our models are the first to be trained on somatic cancer mutation data, they demonstrate proof-of-concept, leverage orthogonal lines of evidence, and warrant consideration for use in aggregator tools. The supervised learning models in our study can be implemented using the training dataset, and subsequently applied to variants of interest prospectively to obtain probability scores of pathogenicity. While the LRM is restricted to the 66 genes constituting our training dataset, the RFM is not limited to these genes. Through our collaborations with MSSNG, C4R, GEL, and GeneDx, we identified an additional 65 individuals with suspected rare diseases and a germline variant that overlapped with a Cancer Hotspot mutation. Many of these cases remain “unsolved”, and the inclusion of this criterion may offer valuable insights for variant interpretation.

This study focused on missense variants because of the existence of a cancer driver missense mutation database and because of the large number of missense variants in ClinVar. We explored the potential application of using cancer missense mutations to inform germline variant interpretation to non-coding variants by leveraging mutation data from COSMIC and other putative cancer driver databases (Supplemental Methods). Results were inconclusive due to the limited availability of non-coding germline variants clinically classified in public databases.

This study has several additional limitations. It primarily focused on a subset of cancer mutations from Cancer Hotspots, last updated in 2017. However, only a small fraction of the additional highly recurrent missense mutations present in COSMIC in 2024 overlapped with germline variants in ClinVar, suggesting that Cancer Hotspots remains a near comprehensive list of statistically recurring cancer (driver) mutations. We did not assess the oncogenicity of each cancer mutation in Cancer Hotspots.^33^ There are 41 tumour types represented in Cancer Hotspots, with the majority being solid tumours in adults.^23^ The inclusion of more tumour tissue types over time will likely result in the identification of additional driver mutations. This study used ClinVar as the set of germline missense variants, and while filtering steps were applied, we acknowledge that the quality of ClinVar entries is not equal. Additionally, it is possible that overlap with cancer mutations contributed to the clinical interpretation of some germline variants in ClinVar, despite such evidence not yet being codified in existing classification guidelines.^4,34,35^ Of note, however, is that the term “Cancer Hotspots database” was only mentioned 3 times in the context of missense SNVs in the ClinVar database of 3,614,935 submitted records (search date: December 2023). In the training dataset, there was variability in the LRM’s independent “gene” variable, leading to inconsistent performance across genes. Future work will focus on conducting gene-level model evaluations once larger datasets become available, providing more statistical power to assess gene-specific effects.^36^ None of the *in silico* prediction tools used in this study address variant pathomechanism (i.e., gain of function, loss of function). We recognize the potential relevance of this consideration, particularly for germline missense variants with a gain of function mechanism, where *in silico* tools like REVEL demonstrate worse performance.^37^ The absence of this consideration may limit the applicability of the findings in cases where different disease mechanisms are at play between cancer mutations and germline variants (e.g., variants in *MYD88*, where germline variants can lead to immunodeficiency through loss of function^38,39^, but acts as a proto-oncogene in cancer^40^). Even when the germline phenotype is cancer-related there may be discrepancies in mechanism (e.g., TERT loss of function in the germline versus increased expression somatically in certain tumours).^41^ Further increasing the size of the test dataset was not possible; to compensate, cross-validation was used to evaluate model performance. Last, while we identified additional germline variants that overlap with CH mutations in private genomic datasets, we were not able to formally reclassify variants and return new information back to those individuals. However, the identified variants in the GEL Research Environment were shared with GEL for further review.

Our results demonstrate a modeling approach that uses overlapping cancer mutations to facilitate the interpretation of pathogenic germline missense variants. The presence of a variant in Cancer Hotspots suggests that additional published evidence from somatic cancer studies exists that may be relevant to understanding the impact of the same variant in a germline context. There are clear definitions of somatic mutational hotspots^33^, that can be applied to future published cancer datasets, enabling better applications of our tool. As we navigate the complexities of variant interpretation, leveraging the growing wealth of genomic data in both cancer and germline contexts will contribute to refining our understanding and improving diagnostic capabilities in the field of rare diseases.

## METHODS

### Extracting cancer mutation data from Cancer Hotspots

We obtained cancer mutation data for 3,122 single nucleotide variants (SNVs) from the Cancer Hotspots^22,23^ database (www.cancerhotspots.org), representing a set of true cancer driver mutations. This database consists of mutational hotspots identified in large scale cancer genomics data, defined as single amino acid positions in protein-coding genes that are mutated more frequently than would be expected in the absence of selection.^13,23^ This method assigns a statistical significance to the recurrence of mutation at a given amino acid and is corrected for background mutational rate of the position, gene, and sample both within and across cancer types in the affected cohort.^22,23^ Somatic mutational hotspots are therefore not common germline benign variants in a population.^13,22,23^ A Python script was developed to extract genomic coordinates in GRCh37, reference and alternate alleles, and tumour sample counts for each mutation. Only missense mutations (n=2,576) were used for our analyses. We annotated the cancer missense mutations using ANNOVAR and a custom pipeline^2^ developed by The Centre for Applied Genomics (Toronto, Canada). ClinVar annotations (date accessed: Jan 2022) were used to identify clinical classifications of those germline variants that are also cancer mutations in Cancer Hotspots. We conservatively excluded any mutations with corresponding germline variants with “conflicting interpretations of pathogenicity” (CIP) or considered a “risk factor” for disease (n = 129). The remaining 2,447 recurrent missense mutations (n=216 total genes) from Cancer Hotspots are hereafter referred to as the “CH mutations”.

### Comparing cancer mutations with germline variants

Separately, we extracted from ClinVar (date accessed: Jan 2022) all missense variants in the 216 genes from the list of CH mutations (n = 51,346 SNVs) (Supplemental Figure 1). We selected missense variants with a “germline” allele origin, i.e., excluding those labeled as “somatic” or “unknown”. These variants were then grouped into three categories based on their ACMG classification in ClinVar: “likely pathogenic” or “pathogenic” (LP/P) (n = 3,149), “likely benign” or “benign” (LB/B) (n = 2,755), and “variant of uncertain significance” (VUS) (n = 45,442). We annotated these variants using ANNOVAR to include REVEL^42^, phyloP^43^ (20way mammalian and 7way vertebrate), and phastCons^44^ (20way mammalian and 7way vertebrate) scores. For each variant, we noted the presence or absence of an overlap with a CH mutation. These variants are hereafter to as the “ClinVar dataset” and were used to calculate the odds ratios of a germline variant that overlaps with a CH mutation having an LP/P classification. This data was also used to apply mathematical framework described by Tavtigian et al. to define ACMG/AMP evidence strength for the use of cancer mutational hotspot data for germline variant interpretation.^27^

### Identifying overlap with cancer mutations in other genomic databases

We queried the CH mutations in four controlled-access GWS databases, in collaboration with MSSNG^45^, Genomics England^46^ (GEL), Care4Rare^47^ (C4R), and GeneDx^9,48^, to identify matching germline missense variants (at the nucleotide level).

The MSSNG database represents a cohort of autistic individuals / individuals with autism and their family members. All germline missense variants in this database were extracted and converted to GRCh37 using LiftOver. Germline variants in MSSNG, and CH mutations, were imported to R version 4.1.0 (R Foundation for Statistical Computing) to identify overlapping variants by genomic coordinate, reference allele, and alternate allele. The GEL, C4R, and GeneDx databases represent phenotypically heterogeneous cohorts of individuals with suspected rare genetic diseases and their family members. In the GEL Research Environment, a bash shell script was used to extract variants from variant call format (VCF) files by genomic coordinates. The CH mutations were queried against germline variants in the VCF files of all participants in the Rare Disease program of GEL using this script. The participant IDs for each CH mutation that overlapped with a germline variant in GEL were used to retrieve phenotype data along with their classifications using the Labkey platform. In collaboration with C4R and GeneDx, the CH mutations were sent to the respective study teams and queried within their databases. Results of overlapping variants and participant IDs were returned. Variant classification and phenotype data from C4R was explored by searching the Genomics4RareDisease (G4RD) database with participant IDs.^49^

### Identifying cancer mutations from other cancer databases and comparing with germline variants

We downloaded approximately 1.1 million coding mutations from the COSMIC database^50^ listed in the Cancer Gene Census^25^ and filtered for confirmed somatic missense mutations (n = 231, 477). To align with the stringent criteria used in the Cancer Hotspots database, we further filtered based on the presence of mutations in COSMIC across a defined number of tumor samples. This step ensured the retention of only those mutations observed across a substantial number of tumors, indicative of potential driver mutations as defined in Cancer Hotspots. For this filtering process, we used tumor sample counts of CH mutations that overlap with germline variants in ClinVar (Supplemental Methods). Plotting these values by ClinVar classification groups (LP/P and LB/B/VUS), we generated receiver operating characteristic (ROC) curves to determine the optimal tumor sample count cut-off for distinguishing between LP/P and LB/B/VUS variants. The identified optimal count was then used to filter the COSMIC mutations. We then conducted further filtered to identify “new” mutations in COSMIC, i.e., those absent in Cancer Hotspots, and compared these mutations with germline variants in ClinVar, to identify additional overlapping variants.

### Training dataset used for supervised learning models

We developed supervised learning models to predict pathogenicity of unclassified germline variants, based on a set of variants with known classifications in ClinVar. To construct the training variant set, we used the ClinVar dataset including n = 51,346 SNVs in the 216 genes from the list of CH mutations. Different nucleotide variants resulting in the same amino acid change were grouped together. VUS with REVEL scores >0.29 were excluded from the training dataset. This cut-off is the upper-most bound for BP4 evidence level for REVEL scores.^28^ The remaining VUS were included and treated as LB/B variants (Figure 2; see below regarding weighting), to address class imbalance arising from fewer LB/B versus LP/P variants in the dataset. Variants were then restricted to a set of 66 genes, determined by the updated list of 428 CH mutations overlapping with germline variants (Figure 2). The resulting training dataset comprises 13,881 variants.

### Developing supervised learning models

Two types of supervised learning models were fit to the training dataset in R: a logistic regression model (LRM) and a random forest model (RFM). Pathogenicity status (LB/B, LP/P) was used as the dependent variable and the following were used as independent variables: 1) overlap with a cancer missense mutation from Cancer Hotspots (2 categories: present = 1, absent = 0), 2) the protein-coding gene associated with a variant (with 66 categories representing each gene), 3) the number of tumour samples with a specific amino acid change at a residue position from Cancer Hotspots, 4) the number of tumour samples with a mutated residue from Cancer Hotspots, 5 & 6) the phyloP conservation scores^43^ (20way mammalian and 7way vertebrate), and 7 & 8) the phastCons conservation scores^44^ (20way mammalian and 7way vertebrate).

The ‘stats’ R package was used to fit the LRM. REVEL scores for the included VUS (all <= 0.29) were used as prior weights (*weight* = 1 - *REVEL score*) compared to true LB/B variants (*weight* = 1). The predicted probabilities and standard performance metrics including Akaike Information Criterion (AIC) and McFadden’s pseudo-R^2^ were used to assess the fit of the model. The same training dataset was used for the RFM using the ‘randomForest’ package in R. However, the gene variable was excluded due to a categorical variable limit of 32 levels. 350 classification trees were generated, and four independent variables were randomly selected as candidates for each split in the classification trees.

### Evaluating supervised learning models with test dataset

Both LRM and RFM performance was evaluated using a test dataset of 332 germline missense variants that were absent from the training dataset. These variants were obtained from new ClinVar submissions from Feb 2022 to Aug 2022 (n = 189), the Leiden Open Variation Database (LOVD)^51^ (n = 35), G4RD database^54^ (n = 1), GEL database^52^ (n = 93), SickKids Cancer Sequencing (KiCS) dataset^53^ (n = 2), and from manual review of literature pertaining to the genes of interest that was published from 2021-2022 (n = 19). The test dataset variants impact genes that are represented in the training dataset. We used the predicted classifications of each model across all possible classification thresholds to plot precision-recall curves and calculate the area under the curve (AUPRC). The highest performing model and optimal threshold were used to assess the pathogenicity of an additional set of variants with unknown classification identified in other genomic databases through collaborations. The variants in the test dataset were annotated using scores from other *in silico* prediction tools, including SIFT^54^, PolyPhen-2^55^, REVEL^42^, CADD^56^, VARITY^57^, AlphaMissense^29^, PrimateAI^10^, VEST4^58^, and MutPred2^59^. Some tools were selected because they are commonly used for variant interpretation in the diagnostic laboratory, are referenced in ACMG/AMP guidelines,^4^ and/or are incorporated into annotation tools like ANNOVAR. The remaining tools (e.g., AlphaMissense) were selected because of their strong potential to be incorporated into clinical interpretation workflows in the future. We also plotted precision-recall curves using these scores to calculate the AUPRCs and compared them with the LRM and RFM.

### Evaluating supervised learning models with cross-validation

Cross-validation was conducted using the ‘caret’ package in R, with the ‘createFolds’ function employed to generate the folds for model training and evaluation. The training dataset was divided into *k* folds, where the model was trained on *k-1* fold and tested on the remaining one. The training dataset was divided into 8 and 10 folds for the LRM and RFM, respectively. The F1 score and AUPRC, using a threshold of 0.5, was calculated for each fold, and averaged over the *k* folds to obtain an estimate of each model’s generalization ability.

### Statistical methods

Standard descriptive statistics, odds ratios, and Mann-Whitney U tests were performed using R and GraphPad Prism 9 with two-tailed statistical significance set at p < 0.05.

## LIST OF ABBREVIATIONS

## DECLARATIONS

## ETHICS DECLARATION

This secondary use data study was approved by the Research Ethics Board at the Hospital for Sick Children. The de-identified data from GeneDx was assessed in accordance with an IRB-approved protocol (WIRB #20171030).

## AVAILABILITY OF DATA AND MATERIALS

The cancer mutation data from Cancer Hotspots that support the findings of this study are available through a public database and at the following URL: https://www.cancerhotspots.org/ (DOI: <u>10.1038/nbt.3391</u>) Germline variants and their classifications are available in the ClinVar public archive: https://www.ncbi.nlm.nih.gov/clinvar/ (DOI: https://doi.org/10.1093/nar/gkx1153). For the Cancer Hotspots cancer mutation data transformation, the Python script is openly available on a GitHub repository: https://github.com/haqueb2/Cancer-Hotspots-Reformat. The training dataset used for training supervised learning models, the LRM and RFM pathogenicity scores assigned to training and test dataset variants, and prediction scores generated by other *in silico* tools for the test dataset are all available in Supplemental Table 6. All variants used in test and training datasets are included in Supplemental Table 6. R scripts used to train supervised learning models can be found in Supplemental Appendix 1 and 2. Datasets from Genomics England (DOI: https://doi.org/10.6084/m9.figshare.4530893.v7), MSSNG (DOI: <u>10.1016/j.cell.2022.10.009</u>), Care4Rare (DOI:<u>10.1016/j.ajhg.2022.10.002</u>), and GeneDx are not openly available due to controlled access requirements. Access to these datasets can be made available upon request to the respective organizations.

## ACKNOWLEDGEMENTS

This research was made possible through access to data in the National Genomic Research Library, which is managed by Genomics England Limited (a wholly owned company of the Department of Health and Social Care). The National Genomic Research Library holds data provided by patients and collected by the NHS as part of their care and data collected as part of their participation in research. The National Genomic Research Library is funded by the National Institute for Health Research and NHS England. The Wellcome Trust, Cancer Research UK and the Medical Research Council have also funded research infrastructure. The authors wish to acknowledge the resources of MSSNG (www.mss.ng), Autism Speaks and The Centre for Applied Genomics at The Hospital for Sick Children, Toronto, Canada. We also thank the participating families for their time and contributions to this database, as well as the generosity of the donors who supported this program. This study makes use of data obtained through Care4Rare Canada studies (CHEO REB #11/04E and OGI-147) and shared via controlled access to Genomics4RD, a rare disease data sharing platform. We are grateful to the biostatisticians through the Clinical Research Core Facilities at the Hospital for Sick Children for their consultation on training data design and statistical analyses. We thank additional students affiliated with the Department of Molecular Genetics at the University of Toronto who provided helpful input on study design and analysis plans.

## AUTHOR CONTRIBUTIONS

Conceptualization: GC

Data curation: BH, TM, BT, TH, MMM, EMP

Formal analysis: BH, DC, AP, MC, JN, CS, JZ

Funding acquisition: BH, GC

Supervision: GC, DM, FPR

Visualization: BH

Writing-original draft: BH, GC

Writing-review & editing: DC, AP, MC, TN, JN, CS, BT, JZ, TH, MMM, EMP, SW, DM, FPR

