## Supplemental for "Leveraging cancer mutation data to predict the pathogenicity of germline missense variants"

**SUPPLEMENTAL FILE FOR:** Leveraging cancer mutation data to predict the pathogenicity of germline missense variants

**TABLE OF CONTENTS**

Supplemental Methods…………………………………………………………………...………2

Supplemental Table 1……………………………………………………………………...….…5

Supplemental Table 2……………………………………………………………………...…....13

Supplemental Table 3……………………………………………………………………...…....17

Supplemental Table 4……………………………………………………………………...…....18

Supplemental Table 5……………………………………………………………………...…....19

Supplemental Table 6…………………………………………………………[separate .xls file]

Supplemental Figure 1…………………………………………………………...……………...20

Supplemental Figure 2…………………………………………………………….……………21

Supplemental Figure 3…………………………………………………………….....…………22

Supplemental Figure 4…………………………………………………………………...……..23

Supplemental Figure 5…………………………………………………………………...……..24

Supplemental Figure 6…………………………………………………………………...……..25

Supplemental Figure 7…………………………………………………………………...……..26

Supplemental Figure 8…………………………………………………………………...……..27

Supplemental Figure 9…………………………………………………………………...……..28

Supplemental Figure 10……..…………………………………………………………...……..29

Supplemental Figure 11……………………………………………………………….....……..30

Supplemental Figure 12……………………………………………………………….....……..31

Supplemental Appendix 1…………………………………………………………...…………..32

Supplemental Appendix 2…………………………………………………………...…………..36

Supplemental References…………………………………………………………...……………38

List of Legends…………………………………………………………...………………...……39

**Supplemental Methods**

***Identifying functional cancer evidence for mutations from Cancer Hotspots***

We searched for mutations in cancer genes sourced from Cancer Hotspots (n = 216) in the Jackson Laboratory Clinical Knowledgebase (CKB) database.^1^ In CKB, the protein-level effect of each mutation is categorized as "loss of function," "gain of function," "no effect," or "unknown", based on manual curation of the published literature. We did not assess whether the functional evidence from CKB aligns with ClinGen recommendations for a "well-established" functional assay to qualify for the PS3 criterion.^2^

***Applying lines of evidence for germline variant classification***

We calculated the positive likelihood ratio (LR+) that assesses the degree to which the overlap of a germline missense variant with a Cancer Hotspot mutation increases the odds that the germline variant is classified as LP/P in ClinVar.^3^ For each VUS in ClinVar that overlapped with Cancer Hotspots, we determined the ACMG/AMP variant classification evidence codes that could be applied in the absence of clinical/family data (e.g., PM2_Supporting, PP2, PP3, PP3_Moderate, and PP3_Strong). The ACMG/AMP combining rules were then used to clarify whether additional evidence from the cancer LR+ could hypothetically upgrade VUS classifications to LP/P.

***Plotting distribution of REVEL and AlphaMissense scores***

The REVEL and AlphaMissense scores were obtained for the ClinVar dataset (pre-filtering).^4,5^ We plotted the distribution of both scores for germline variants in ClinVar that overlapped with cancer mutations in Cancer Hotspots (CH+, ClinVar+), germline variants that do not overlap with cancer mutations (CH-, ClinVar+), and all remaining cancer mutations that do not overlap with germline variants (CH+, ClinVar-). The median score was calculated for each group.

***Exploratory analysis with independent variables prior to model training***

We performed an analysis of all independent variables to evaluate their overall impact on the ClinVar dataset (pre-filtering). Specifically, we focused on a subset of eight genes (*TP53*, *PIK3CA*, *PTEN*, *SMAD4*, *VHL*, *PTPN11*, *RIT1*, and *FGFR3*), plotting their total number of cancer mutations from Cancer Hotspots. The inclusion of the proportion of these mutations overlapping with germline variants in ClinVar allowed us to discern variations in LP/P variant enrichment across these genes, highlighting their relative pathogenicity within the model.

We also evaluated the ability of tumor sample counts from CH cancer mutations overlapping with ClinVar germline variants to differentiate between LP/P and LB/B/VUS classifications. Plotting these values by ClinVar classification groups (LP/P and LB/B/VUS), we generated receiver operating characteristic (ROC) curves and compared area under the curve (AUC) values to determine the tumor sample count cut-off that best discriminates between the two groups of classifications. We found this cut-off to be >25 tumour sample counts, which also influences predicted pathogenicity scores for the LRM, and represents a cut-off for a node in RFM decision trees by splitting the training data with the lowest Gini impurity.

Additionally, we generated four plots to visualize phyloP and phastCons scores (both mammalian and vertebrate scores). Each plot displayed the distribution of LP/P and VUS variants for the CH+, ClinVar+ and CH-, ClinVar+ variant groups. We compared these variant groups (CH+, ClinVar+ versus CH-, ClinVar+) by median scores and computing the probability of superiority (PS), to evaluate the relative superiority of values between the two groups.

***Investigating overlap of non-coding cancer mutations from COSMIC with germline variants***

Non-coding cancer mutations (n = 434,213) in GRCh37 were downloaded from the COSMIC database^6^ (release date: May 2023) and annotated using a custom pipeline developed by The Centre for Applied Genomics (TCAG) in Toronto, Canada. ClinVar annotations (date accessed: Sep 2023) were used to identify mutations that were observed as germline variants along with their classifications. However, we observed a modest degree of overlap and we were underpowered to employ similar methods as for the missense mutations from Cancer Hotspots (data not shown). In many cases, ClinVar classifications for variants corresponded to the coding region of an alternate transcript, rather than the non-coding region where cancer mutation was observed.

Supplemental Table 1. Genes with missense mutations from Cancer Hotspots that overlap with germline variants in ClinVar, along with their OMIM disease phenotype(s).

| **Gene** | **Cancer Gene Type** | **Phenotype Classification** | **OMIM Disease Phenotype** |
| --- | --- | --- | --- |
| ACVR1 | Proto-oncogene | Multisystem phenotype | Fibrodysplasia ossificans progressiva, 135100 (3), i:AD |
| AKT1 | Proto-oncogene | Multisystem phenotype | Breast cancer, somatic, 114480 (3); Cowden syndrome 6, 615109 (3); Proteus syndrome, somatic, 176920 (3); Ovarian cancer, somatic, 167000 (3); Colorectal cancer, somatic, 114500 (3) |
| ALK | Oncogene, fusion | Hereditary cancer predisposition phenotype | {Neuroblastoma, susceptibility to, 3}, 613014 (3) |
| APC | Tumour suppressor gene | Hereditary cancer predisposition phenotype | Adenoma, periampullary, somatic, 175100 (3); Desmoid disease, hereditary, 135290 (3), i:AD; Adenomatous polyposis coli, 175100 (3), i:AD; Gardner syndrome, 175100 (3), i:AD; Hepatoblastoma, somatic, 114550 (3); Colorectal cancer, somatic, 114500 (3); Gastric adenocarcinoma and proximal polyposis of the stomach, 619182 (3), i:AD; Brain tumor-polyposis syndrome 2, 175100 (3), i:AD; Gastric cancer, somatic, 613659 (3) |
| ATM | Tumour suppressor gene | Multisystem phenotype/Hereditary cancer predisposition phenotype | Lymphoma, mantle cell, somatic (3); Lymphoma, B-cell non-Hodgkin, somatic (3); Ataxia-telangiectasia, 208900 (3), i:AR; {Breast cancer, susceptibility to}, 114480 (3), i:AD, i:SMu; T-cell prolymphocytic leukemia, somatic (3) |
| BRAF | Oncogene, fusion | Multisystem phenotype | Nonsmall cell lung cancer, somatic (3); Noonan syndrome 7, 613706 (3), i:AD; Cardiofaciocutaneous syndrome, 115150 (3), i:AD; Colorectal cancer, somatic, 114500 (3); Adenocarcinoma of lung, somatic, 211980 (3); Melanoma, malignant, somatic,, 155600 (3); LEOPARD syndrome 3, 613707 (3), i:AD |
| BRCA2 | Tumour suppressor gene | Hereditary cancer predisposition phenotype | {Pancreatic cancer 2}, 613347 (3); {Breast cancer, male, susceptibility to}, 114480 (3), i:AD, i:SMu; {Glioblastoma 3}, 613029 (3), i:AR; Wilms tumor, 194070 (3), i:AD, i:SMu; Fanconi anemia, complementation group D1, 605724 (3), i:AR; {Medulloblastoma}, 155255 (3), i:AD, i:AR, i:SMu; {Prostate cancer}, 176807 (3), i:AD, i:SMu; {Breast-ovarian cancer, familial, 2}, 612555 (3), i:AD |
| CARD11 | Proto-oncogene | Multisystem phenotype | Immunodeficiency 11B with atopic dermatitis, 617638 (3), i:AD; B-cell expansion with NFKB and T-cell anergy, 616452 (3), i:AD; Immunodeficiency 11A, 615206 (3), i:AR |
| CASP8 | Tumour suppressor gene | Multisystem phenotype | {Lung cancer, protection against}, 211980 (3), i:AD, i:SMu; ?Autoimmune lymphoproliferative syndrome, type IIB, 607271 (3), i:AR; Hepatocellular carcinoma, somatic, 114550 (3); {Breast cancer, protection against}, 114480 (3), i:AD, i:SMu |
| CDH1 | Tumour suppressor gene | Multisystem phenotype/Hereditary cancer predisposition phenotype | Endometrial carcinoma, somatic, 608089 (3); {Prostate cancer, susceptibility to}, 176807 (3), i:AD, i:SMu; Blepharocheilodontic syndrome 1, 119580 (3), i:AD; Gastric cancer, hereditary diffuse, with or without cleft lip and/or palate, 137215 (3), i:AD; {Breast cancer, lobular}, 114480 (3), i:AD, i:SMu; Ovarian cancer, somatic, 167000 (3) |
| CDK4 | Proto-oncogene | Hereditary cancer predisposition phenotype | {Melanoma, cutaneous malignant, 3}, 609048 (3), i:AD |
| CDKN1B | Tumour suppressor gene | Hereditary cancer predisposition phenotype | Multiple endocrine neoplasia, type IV, 610755 (3), i:AD |
| CDKN2A | Tumour suppressor gene | Hereditary cancer predisposition phenotype | {Melanoma and neural system tumor syndrome}, 155755 (3), i:AD; {Melanoma, cutaneous malignant, 2}, 155601 (3), i:AD; {Melanoma-pancreatic cancer syndrome}, 606719 (3), i:AD |
| CHEK2 | Tumour suppressor gene | Hereditary cancer predisposition phenotype | Li-Fraumeni syndrome, 609265 (3); Osteosarcoma, somatic, 259500 (3); {Prostate cancer, familial, susceptibility to}, 176807 (3), i:AD, i:SMu; {Breast cancer, susceptibility to}, 114480 (3), i:AD, i:SMu; {Breast and colorectal cancer, susceptibility to} (3) |
| CREBBP | oncogene, TSG, fusion | Multisystem phenotype | Rubinstein-Taybi syndrome 1, 180849 (3), i:AD; Menke-Hennekam syndrome 1, 618332 (3), i:AD |
| CTCF | Tumour suppressor gene | Multisystem phenotype | Mental retardation, i:AD 21, 615502 (3), i:AD |
| CTLA4 | Oncogene(?) | Multisystem phenotype | {Systemic lupus erythematosus, susceptibility to}, 152700 (3), i:AD; {Diabetes mellitus, insulin-dependent, 12}, 601388 (3); {Celiac disease, susceptibility to, 3}, 609755 (3); Autoimmune lymphoproliferative syndrome, type V, 616100 (3), i:AD; {Hashimoto thyroiditis}, 140300 (3), i:AD |
| CTNNB1 | Oncogene, fusion | Multisystem phenotype | Ovarian cancer, somatic, 167000 (3); Colorectal cancer, somatic, 114500 (3); Medulloblastoma, somatic, 155255 (3); Hepatocellular carcinoma, somatic, 114550 (3); Pilomatricoma, somatic, 132600 (3); Neurodevelopmental disorder with spastic diplegia and visual defects, 615075 (3), i:AD; Exudative vitreoretinopathy 7, 617572 (3), i:AD |
| DICER1 | Tumour suppressor gene | Hereditary cancer predisposition phenotype | GLOW syndrome, somatic mosaic, 618272 (3); Rhabdomyosarcoma, embryonal, 2, 180295 (3); Goiter, multinodular 1, with or without Sertoli-Leydig cell tumors, 138800 (3), i:AD; Pleuropulmonary blastoma, 601200 (3), i:AD |
| DNMT1 | N/A | Multisystem phenotype | Cerebellar ataxia, deafness, and narcolepsy, i:AD, 604121 (3), i:AD; Neuropathy, hereditary sensory, type IE, 614116 (3), i:AD |
| DNMT3A | Tumour suppressor gene | Multisystem phenotype | Heyn-Sproul-Jackson syndrome, 618724 (3), i:AD; Acute myeloid leukemia, somatic, 601626 (3); Tatton-Brown-Rahman syndrome, 615879 (3), i:AD |
| EGFR | Proto-oncogene | Multisystem phenotype | ?Inflammatory skin and bowel disease, neonatal, 2, 616069 (3), i:AR; Nonsmall cell lung cancer, response to tyrosine kinase inhibitor in, 211980 (3), i:AD, i:SMu; Adenocarcinoma of lung, response to tyrosine kinase inhibitor in, 211980 (3), i:AD, i:SMu; {Nonsmall cell lung cancer, susceptibility to}, 211980 (3), i:AD, i:SMu |
| ERBB2 | Oncogene, fusion | No OMIM Mendelian disorder | Glioblastoma, somatic, 137800 (3); Adenocarcinoma of lung, somatic, 211980 (3); Gastric cancer, somatic, 613659 (3); Ovarian cancer, somatic (3) |
| ETV6 | Tumour suppressor gene, fusion | Multisystem phenotype | Leukemia, acute myeloid, somatic, 601626 (3); Thrombocytopenia 5, 616216 (3), i:AD |
| FBXW7 | Tumour suppressor gene | Multisystem phenotype | [common variants in Wilms tumor; neurodevelopmental phenotype with variable features, hypotonia and constipation] |
| FGFR1 | Oncogene, fusion | Multisystem phenotype | Pfeiffer syndrome, 101600 (3), i:AD; Jackson-Weiss syndrome, 123150 (3), i:AD; Trigonocephaly 1, 190440 (3), i:AD; Hypogonadotropic hypogonadism 2 with or without anosmia, 147950 (3), i:AD; Hartsfield syndrome, 615465 (3), i:AD; Osteoglophonic dysplasia, 166250 (3), i:AD; Encephalocraniocutaneous lipomatosis, somatic mosaic, 613001 (3) |
| FGFR2 | Oncogene, fusion | Multisystem phenotype | Apert syndrome, 101200 (3), i:AD; Craniosynostosis, nonspecific (3); Jackson-Weiss syndrome, 123150 (3), i:AD; Scaphocephaly and Axenfeld-Rieger anomaly (3); Saethre-Chotzen syndrome, 101400 (3), i:AD; Gastric cancer, somatic, 613659 (3); Scaphocephaly, maxillary retrusion, and mental retardation, 609579 (3); Bent bone dysplasia syndrome, 614592 (3), i:AD; LADD syndrome, 149730 (3), i:AD; Craniofacial-skeletal-dermatologic dysplasia, 101600 (3), i:AD; Pfeiffer syndrome, 101600 (3), i:AD; Crouzon syndrome, 123500 (3), i:AD; Beare-Stevenson cutis gyrata syndrome, 123790 (3), i:AD; Antley-Bixler syndrome without genital anomalies or disordered steroidogenesis, 207410 (3), i:AD |
| FGFR3 | Oncogene, fusion | Multisystem phenotype | Muenke syndrome, 602849 (3), i:AD; Nevus, epidermal, somatic, 162900 (3); Thanatophoric dysplasia, type II, 187601 (3), i:AD; Bladder cancer, somatic, 109800 (3); CATSHL syndrome, 610474 (3), i:AD, i:AR; Crouzon syndrome with acanthosis nigricans, 612247 (3), i:AD; Hypochondroplasia, 146000 (3), i:AD; LADD syndrome, 149730 (3), i:AD; Achondroplasia, 100800 (3), i:AD; Thanatophoric dysplasia, type I, 187600 (3), i:AD; Colorectal cancer, somatic, 114500 (3); Spermatocytic seminoma, somatic, 273300 (3); Cervical cancer, somatic, 603956 (3); SADDAN, 616482 (3), i:AD |
| FGFR4 | Proto-oncogene | Multisystem phenotype/Hereditary cancer predisposition phenotype | {Cancer progression/metastasis} (3) |
| FOXP1 | Oncogene, fusion | Multisystem phenotype | Mental retardation with language impairment and with or without autistic features, 613670 (3), i:AD |
| GATA2 | Proto-oncogene | Multisystem phenotype/Hereditary cancer predisposition phenotype | Emberger syndrome, 614038 (3), i:AD; {Myelodysplastic syndrome, susceptibility to}, 614286 (3); Immunodeficiency 21, 614172 (3), i:AD; {Leukemia, acute myeloid, susceptibility to}, 601626 (3), i:AD, i:SMu |
| GNAQ | Proto-oncogene | Multisystem phenotype | Sturge-Weber syndrome, somatic, mosaic, 185300 (3); Capillary malformations, congenital, 1, somatic, mosaic, 163000 (3) |
| GNAS | Proto-oncogene | Multisystem phenotype | ACTH-independent macronodular adrenal hyperplasia, 219080 (3), i:SMu; Pseudohypoparathyroidism Ic, 612462 (3), i:AD; Pseudohypoparathyroidism Ib, 603233 (3), i:AD; Pseudopseudohypoparathyroidism, 612463 (3), i:AD; McCune-Albright syndrome, somatic, mosaic, 174800 (3); Osseous heteroplasia, progressive, 166350 (3), i:AD; Pituitary adenoma 3, multiple types, somatic, 617686 (3); Pseudohypoparathyroidism Ia, 103580 (3), i:AD |
| HNF1A | Tumour suppressor gene | Multisystem phenotype | {Diabetes mellitus, insulin-dependent}, 222100 (3), i:AR; MODY, type III, 600496 (3), i:AD; Hepatic adenoma, somatic, 142330 (3); Renal cell carcinoma, 144700 (3); Diabetes mellitus, insulin-dependent, 20, 612520 (3); {Diabetes mellitus, noninsulin-dependent, 2}, 125853 (3), i:AD |
| HRAS | Proto-oncogene | Multisystem phenotype | Nevus sebaceous or woolly hair nevus, somatic, 162900 (3); Congenital myopathy with excess of muscle spindles, 218040 (3), i:AD; Bladder cancer, somatic, 109800 (3); Thyroid carcinoma, follicular, somatic, 188470 (3); Schimmelpenning-Feuerstein-Mims syndrome, somatic mosaic, 163200 (3); Spitz nevus or nevus spilus, somatic, 137550 (3); Costello syndrome, 218040 (3), i:AD |
| IDH1 | Proto-oncogene | Multisystem phenotype | {Glioma, susceptibility to, somatic}, 137800 (3) |
| IDH2 | Proto-oncogene | Multisystem phenotype | D-2-hydroxyglutaric aciduria 2, 613657 (3) |
| IL7R | Proto-oncogene | Multisystem phenotype | Severe combined immunodeficiency, T-cell negative, B-cell/natural killer cell-positive type, 608971 (3), i:AR |
| KDR | Proto-oncogene | Multisystem phenotype | {Hemangioma, capillary infantile, susceptibility to}, 602089 (3), i:AD; Hemangioma, capillary infantile, somatic, 602089 (3) |
| KIT | Proto-oncogene | Hereditary cancer predisposition phenotype | Gastrointestinal stromal tumor, familial, 606764 (3), i:AD, Isolated cases; Mastocytosis, cutaneous, 154800 (3), i:AD; Germ cell tumors, somatic, 273300 (3); Leukemia, acute myeloid, somatic, 601626 (3); Mastocytosis, systemic, somatic, 154800 (3); Piebaldism, 172800 (3), i:AD |
| KMT2D | Oncogene, tumour suppressor gene | Multisystem phenotype | Kabuki syndrome 1, 147920 (3), i:AD |
| KRAS | Proto-oncogene | Multisystem phenotype | Oculoectodermal syndrome, somatic, 600268 (3); Leukemia, acute myeloid, somatic, 601626 (3); Breast cancer, somatic, 114480 (3); RAS-associated autoimmune leukoproliferative disorder, 614470 (3), i:AD; Cardiofaciocutaneous syndrome 2, 615278 (3), i:AD; Arteriovenous malformation of the brain, somatic, 108010 (3); Bladder cancer, somatic, 109800 (3); Pancreatic carcinoma, somatic, 260350 (3); Lung cancer, somatic, 211980 (3); Gastric cancer, somatic, 137215 (3); Schimmelpenning-Feuerstein-Mims syndrome, somatic mosaic, 163200 (3); Noonan syndrome 3, 609942 (3), i:AD |
| MAP2K1 | Proto-oncogene | Multisystem phenotype | Cardiofaciocutaneous syndrome 3, 615279 (3), i:AD; Melorheostosis, isolated, somatic mosaic, 155950 (3) |
| MAP2K2 | Proto-oncogene | Multisystem phenotype | Cardiofaciocutaneous syndrome 4, 615280 (3), i:AD |
| MAX | Tumour suppressor gene | Hereditary cancer predisposition phenotype | {Pheochromocytoma, susceptibility to}, 171300 (3), i:AD |
| MET | Proto-oncogene | Hereditary cancer predisposition phenotype | {Osteofibrous dysplasia, susceptibility to}, 607278 (3), i:AD; Hepatocellular carcinoma, childhood type, somatic, 114550 (3); ?Deafness, autosomal recessive 97, 616705 (3), i:AR; Renal cell carcinoma, papillary, 1, familial and somatic, 605074 (3) |
| MTOR | Proto-oncogene | Multisystem phenotype | Smith-Kingsmore syndrome, 616638 (3), i:AD; Focal cortical dysplasia, type II, somatic, 607341 (3) |
| MYD88 | Proto-oncogene | Multisystem phenotype | Immunodeficiency 68, 612260 (3), i:AR; Macroglobulinemia, Waldenstrom, somatic, 153600 (3) |
| NF1 | Tumour suppressor gene, fusion | Multisystem phenotype/Hereditary cancer predisposition phenotype | Neurofibromatosis-Noonan syndrome, 601321 (3), i:AD; Leukemia, juvenile myelomonocytic, 607785 (3), i:AD, i:SMu; Neurofibromatosis, familial spinal, 162210 (3), i:AD; Watson syndrome, 193520 (3), i:AD; Neurofibromatosis, type 1, 162200 (3), i:AD |
| NFE2L2 | Oncogene, tumour suppressor gene | Multisystem phenotype | Immunodeficiency, developmental delay, and hypohomocysteinemia, 617744 (3), i:AD |
| NOTCH1 | oncogene, TSG, fusion | Multisystem phenotype | Aortic valve disease 1, 109730 (3), i:AD; Adams-Oliver syndrome 5, 616028 (3), i:AD |
| NRAS | Proto-oncogene | Multisystem phenotype | Epidermal nevus, somatic, 162900 (3); Melanocytic nevus syndrome, congenital, somatic, 137550 (3); Schimmelpenning-Feuerstein-Mims syndrome, somatic mosaic, 163200 (3); Colorectal cancer, somatic, 114500 (3); ?RAS-associated autoimmune lymphoproliferative syndrome type IV, somatic, 614470 (3); Thyroid carcinoma, follicular, somatic, 188470 (3); Neurocutaneous melanosis, somatic, 249400 (3); Noonan syndrome 6, 613224 (3), i:AD |
| PIK3CA | Proto-oncogene | Multisystem phenotype | Ovarian cancer, somatic, 167000 (3); Colorectal cancer, somatic, 114500 (3); CLAPO syndrome, somatic, 613089 (3); Cowden syndrome 5, 615108 (3); Hepatocellular carcinoma, somatic, 114550 (3); Breast cancer, somatic, 114480 (3); Macrodactyly, somatic, 155500 (3); Keratosis, seborrheic, somatic, 182000 (3); Gastric cancer, somatic, 613659 (3); Megalencephaly-capillary malformation-polymicrogyria syndrome, somatic, 602501 (3); Nevus, epidermal, somatic, 162900 (3); CLOVE syndrome, somatic, 612918 (3); Nonsmall cell lung cancer, somatic, 211980 (3) |
| PIK3CD | N/A | Multisystem phenotype | Immunodeficiency 14, 615513 (3), i:AD |
| PIK3R2 | Proto-oncogene | Multisystem phenotype | Megalencephaly-polymicrogyria-polydactyly-hydrocephalus syndrome 1, 603387 (3), i:AD |
| PMS2 | Tumour suppressor gene | Hereditary cancer predisposition phenotype | Colorectal cancer, hereditary nonpolyposis, type 4, 614337 (3); Mismatch repair cancer syndrome 4, 619101 (3) |
| PPP2R1A | Tumour suppressor gene | Multisystem phenotype | Mental retardation, i:AD 36, 616362 (3), i:AD |
| PTEN | Tumour suppressor gene | Multisystem phenotype/Hereditary cancer predisposition phenotype | Prostate cancer, somatic, 176807 (3); {Glioma susceptibility 2}, 613028 (3); Cowden syndrome 1, 158350 (3), i:AD; Lhermitte-Duclos syndrome, 158350 (3), i:AD; Macrocephaly/autism syndrome, 605309 (3), i:AD; {Meningioma}, 607174 (3), i:AD |
| PTPN11 | Proto-oncogene | Multisystem phenotype/Hereditary cancer predisposition phenotype | LEOPARD syndrome 1, 151100 (3), i:AD; Metachondromatosis, 156250 (3), i:AD; Noonan syndrome 1, 163950 (3), i:AD; Leukemia, juvenile myelomonocytic, somatic, 607785 (3) |
| RAC1 | Proto-oncogene | Multisystem phenotype | Mental retardation, i:AD 48, 617751 (3), i:AD |
| RAD50 | Tumour suppressor gene | Multisystem phenotype/Hereditary cancer predisposition phenotype | Nijmegen breakage syndrome-like disorder, 613078 (3) |
| RAD51C | Fusion | Hereditary cancer predisposition phenotype | {Breast-ovarian cancer, familial, susceptibility to, 3}, 613399 (3); Fanconi anemia, complementation group O, 613390 (3), i:AR |
| RAF1 | Oncogene, fusion | Multisystem phenotype | LEOPARD syndrome 2, 611554 (3); Noonan syndrome 5, 611553 (3), i:AD; Cardiomyopathy, dilated, 1NN, 615916 (3), i:AD |
| RB1 | Tumour suppressor gene | Hereditary cancer predisposition phenotype | Small cell cancer of the lung, somatic, 182280 (3); Bladder cancer, somatic, 109800 (3); Retinoblastoma, trilateral, 180200 (3), i:AD, i:SMu; Osteosarcoma, somatic, 259500 (3); Retinoblastoma, 180200 (3), i:AD, i:SMu |
| RET | Oncogene, fusion | Multisystem phenotype | Multiple endocrine neoplasia IIB, 162300 (3), i:AD; Pheochromocytoma, 171300 (3), i:AD; Multiple endocrine neoplasia IIA, 171400 (3), i:AD; Medullary thyroid carcinoma, 155240 (3), i:AD; {Hirschsprung disease, protection against}, 142623 (3), i:AD; Central hypoventilation syndrome, congenital, 209880 (3), i:AD; {Hirschsprung disease, susceptibility to, 1}, 142623 (3), i:AD |
| RHOA | Oncogene, tumour suppressor gene | Multisystem phenotype | Ectodermal dysplasia with facial dysmorphism and acral, ocular, and brain anomalies, somatic mosaic, 618727 (3) |
| RIT1 | Proto-oncogene | Multisystem phenotype | Noonan syndrome 8, 615355 (3), i:AD |
| RRAS2 | Proto-oncogene | Multisystem phenotype | Ovarian carcinoma (3); Noonan syndrome 12, 618624 (3), i:AD |
| RUNX1 | oncogene, TSG, fusion | Hereditary cancer predisposition phenotype | Platelet disorder, familial, with associated myeloid malignancy, 601399 (3), i:AD; Leukemia, acute myeloid, 601626 (3), i:AD, i:SMu |
| SDHA | Tumour suppressor gene | Hereditary cancer predisposition phenotype | i:MT complex II deficiency, nuclear type 1, 252011 (3), i:AR; Cardiomyopathy, dilated, 1GG, 613642 (3), i:AR; Paragangliomas 5, 614165 (3), i:AD |
| SDHAF2 | Tumour suppressor gene | Hereditary cancer predisposition phenotype | Paragangliomas 2, 601650 (3), i:AD |
| SETD2 | Tumour suppressor gene | Multisystem phenotype | Luscan-Lumish syndrome, 616831 (3), i:AD |
| SMAD3 | Tumour suppressor gene | Multisystem phenotype | Loeys-Dietz syndrome 3, 613795 (3), i:AD |
| SMAD4 | Tumour suppressor gene | Multisystem phenotype/Hereditary cancer predisposition phenotype | Polyposis, juvenile intestinal, 174900 (3), i:AD; Juvenile polyposis/hereditary hemorrhagic telangiectasia syndrome, 175050 (3), i:AD; Myhre syndrome, 139210 (3), i:AD; Pancreatic cancer, somatic, 260350 (3) |
| SMARCA4 | Tumour suppressor gene | Hereditary cancer predisposition phenotype | {Rhabdoid tumor predisposition syndrome 2}, 613325 (3), i:AD; Coffin-Siris syndrome 4, 614609 (3), i:AD |
| SMARCB1 | Tumour suppressor gene | Multisystem phenotype/Hereditary cancer predisposition phenotype | Rhabdoid tumors, somatic, 609322 (3); {Schwannomatosis-1, susceptibility to}, 162091 (3), i:AD; Coffin-Siris syndrome 3, 614608 (3), i:AD; {Rhabdoid tumor predisposition syndrome 1}, 609322 (3), i:AD |
| SMO | Proto-oncogene | Multisystem phenotype | Curry-Jones syndrome, somatic mosaic, 601707 (3); Pallister-Hall-like syndrome, 241800 (3), i:AR; Basal cell carcinoma, somatic, 605462 (3) |
| STAT3 | Proto-oncogene | Multisystem phenotype | Hyper-IgE recurrent infection syndrome, 147060 (3), i:AD; Autoimmune disease, multisystem, infantile-onset, 1, 615952 (3), i:AD |
| STK11 | Tumour suppressor gene | Hereditary cancer predisposition phenotype | Testicular tumor, somatic, 273300 (3); Peutz-Jeghers syndrome, 175200 (3), i:AD; Melanoma, malignant, somatic, 155600 (3); Pancreatic cancer, somatic, 260350 (3) |
| TGFBR1 | Oncogene(?) | Multisystem phenotype | Loeys-Dietz syndrome 1, 609192 (3), i:AD; {Multiple self-healing squamous epithelioma, susceptibility to}, 132800 (3), i:AD |
| TGFBR2 | Tumour suppressor gene | Multisystem phenotype/Hereditary cancer predisposition phenotype | Esophageal cancer, somatic, 133239 (3); Colorectal cancer, hereditary nonpolyposis, type 6, 614331 (3); Loeys-Dietz syndrome 2, 610168 (3), i:AD |
| TP53 | oncogene, TSG, fusion | Multisystem phenotype/Hereditary cancer predisposition phenotype | {Adrenocortical carcinoma, pediatric}, 202300 (3), i:AD; {Glioma susceptibility 1}, 137800 (3), i:AD, i:SMu; {Basal cell carcinoma 7}, 614740 (3), i:AD; Bone marrow failure syndrome 5, 618165 (3), i:AD; {Colorectal cancer}, 114500 (3), i:AD, i:SMu; Nasopharyngeal carcinoma, somatic, 607107 (3); Breast cancer, somatic, 114480 (3); {Osteosarcoma}, 259500 (3), i:SMu; {Choroid plexus papilloma}, 260500 (3), i:AD; Li-Fraumeni syndrome, 151623 (3), i:AD; Hepatocellular carcinoma, somatic, 114550 (3); Pancreatic cancer, somatic, 260350 (3) |
| TP63 | Oncogene, tumour suppressor gene | Multisystem phenotype | Limb-mammary syndrome, 603543 (3), i:AD; Orofacial cleft 8, 618149 (3); Split-hand/foot malformation 4, 605289 (3), i:AD; Hay-Wells syndrome, 106260 (3), i:AD; Ectrodactyly, ectodermal dysplasia, and cleft lip/palate syndrome 3, 604292 (3), i:AD; Rapp-Hodgkin syndrome, 129400 (3), i:AD; ADULT syndrome, 103285 (3), i:AD |
| VHL | Tumour suppressor gene | Hereditary cancer predisposition phenotype | Pheochromocytoma, 171300 (3), i:AD; Erythrocytosis, familial, 2, 263400 (3), i:AR; von Hippel-Lindau syndrome, 193300 (3), i:AD; Renal cell carcinoma, somatic, 144700 (3); Hemangioblastoma, cerebellar, somatic (3) |

**Supplemental Table 2.** Genes from Cancer Hotspots with known modes of inheritance for associated Mendelian disease(s), including somatic and germline mechanisms of action and concordance status (concordant, discordant, or semi-concordant).

| **Gene** | **Cancer Mechanism** | **Mendelian Mechanism** | **Concordance** |
| --- | --- | --- | --- |
| ACVR1 | GoF | GoF | Concordant |
| AKT1 | GoF | GoF | Concordant |
| AKT3 | GoF | GoF, LoF | Semi-concordant |
| ALK | GoF | GoF | Concordant |
| ANKRD11 | LoF | LoF | Concordant |
| APC | LoF | LoF | Concordant |
| AR | GoF | GoF | Concordant |
| ARAF | GoF | GoF | Concordant |
| ARID1A | LoF | LoF | Concordant |
| ARID1B | LoF | LoF | Concordant |
| ARID2 | LoF | LoF | Concordant |
| ASXL2 | LoF | LoF | Concordant |
| ATM | LoF | LoF | Concordant |
| AXL | LoF | LoF | Concordant |
| BCL10 | LoF | LoF | Concordant |
| BCL2 | GoF | GoF, LoF | Semi-concordant |
| BCL2L11 | GoF | GoF | Concordant |
| BCOR | LoF | LoF | Concordant |
| BRAF | GoF | GoF | Concordant |
| BRCA2 | LoF | LoF | Concordant |
| BRD4 | GoF | LoF | Discordant |
| CARD11 | GoF | GoF, LoF | Semi-concordant |
| CASP8 | LoF | LoF | Concordant |
| CBL | GoF, LoF | GoF | Semi-concordant |
| CDH1 | LoF | LoF | Concordant |
| CDK12 | LoF | LoF | Concordant |
| CDK4 | GoF | GoF | Concordant |
| CDKN1B | LoF | LoF | Concordant |
| CDKN2A | LoF | LoF | Concordant |
| CHEK2 | LoF | Lof | Concordant |
| CIC | GoF, LoF | LoF | Semi-concordant |
| CREBBP | GoF, LoF | GoF, LoF | Concordant |
| CRLF2 | GoF | GoF | Concordant |
| CTCF | LoF | LoF | Concordant |
| CTLA4 | GoF | LoF | Discordant |
| CTNNB1 | GoF | LoF | Discordant |
| CUL3 | LoF | LoF | Concordant |
| CYSLTR2 | GoF | GoF | Concordant |
| DICER1 | LoF | LoF | Concordant |
| DNMT3A | LoF | GoF, LoF | Semi-concordant |
| DNMT3B | LoF | LoF | Concordant |
| EGFR | GoF | LoF | Discordant |
| EIF1AX | GoF | GoF | Concordant |
| EP300 | LoF | LoF | Concordant |
| EPHA7 | LoF | LoF | Concordant |
| ERBB2 | GoF | LoF | Discordant |
| ERBB3 | GoF | LoF | Discordant |
| ERBB4 | GoF, LoF | GoF, LoF | Concordant |
| ERRFI1 | LoF | LoF | Concordant |
| ESR1 | GoF, LoF | LoF | Semi-concordant |
| ETV6 | LoF | LoF | Concordant |
| EZH2 | GoF, LoF | GoF, LoF | Concordant |
| FAT1 | LoF | LoF | Concordant |
| FGFR1 | GoF | LoF | Discordant |
| FGFR2 | GoF | GoF | Concordant |
| FGFR3 | GoF | GoF, LoF | Semi-concordant |
| FGFR4 | GoF | GoF | Concordant |
| FH | LoF | LoF | Concordant |
| FOXL2 | GoF, LoF | LoF | Semi-concordant |
| FOXP1 | GoF | LoF | Discordant |
| FUBP1 | GoF | LoF | Discordant |
| GATA2 | GoF | LoF | Discordant |
| GATA3 | GoF, LoF | LoF | Semi-concordant |
| GLI1 | GoF | LoF | Discordant |
| GNA11 | GoF | GoF | Concordant |
| GNAS | GoF | LoF, GoF | Semi-concordant |
| GTF2I | GoF | GoF | Concordant |
| H3-3A | GoF | LoF | Discordant |
| H3C2 | GoF | GoF | Concordant |
| HNF1A | LoF | LoF | Concordant |
| HRAS | GoF | GoF | Concordant |
| IDH2 | GoF | GoF | Concordant |
| IKZF1 | LoF | LoF | Concordant |
| IL7R | GoF | LoF | Discordant |
| INPPL1 | LoF | LoF | Concordant |
| JAK1 | GoF, LoF | GoF, LoF | Concordant |
| KDM6A | GoF, LoF | LoF | Semi-concordant |
| KDR | GoF | LoF | Discordant |
| KEAP1 | LoF | LoF | Concordant |
| KIT | GoF | GoF, LoF | Semi-concordant |
| KLF4 | GoF, LoF | GoF, LoF | Concordant |
| KMT2C | LoF | LoF | Concordant |
| KMT2D | GoF, LoF | GoF, LoF | Concordant |
| KNSTRN | GoF | LoF | Discordant |
| KRAS | GoF | GoF | Concordant |
| MAP2K1 | GoF | GoF | Concordant |
| MAP2K2 | GoF | GoF | Concordant |
| MAP3K1 | GoF, LoF | GoF, LoF | Concordant |
| MAPK1 | GoF | GoF | Concordant |
| MED12 | LoF | LoF | Concordant |
| MST1 | LoF | GoF | Discordant |
| MTOR | GoF | GoF | Concordant |
| MYCN | GoF | LoF | Discordant |
| MYD88 | GoF | LoF | Discordant |
| MYOD1 | GoF | LoF | Discordant |
| NF1 | LoF | LoF | Concordant |
| NFE2L2 | GoF, LoF | GoF, LoF | Concordant |
| NOTCH1 | GoF, LoF | GoF, LoF | Concordant |
| NOTCH2 | GoF, LoF | GoF, LoF | Concordant |
| NRAS | GoF | GoF | Concordant |
| PAX5 | GoF, LoF | LoF | Semi-concordant |
| PIK3CA | GoF | GoF | Concordant |
| PIK3R1 | LoF | GoF | Discordant |
| PIK3R2 | GoF | GoF | Concordant |
| PIM1 | GoF | GoF | Concordant |
| PMS2 | LoF | LoF | Concordant |
| POLE | LoF | LoF | Concordant |
| PPP2R1A | LoF | GoF | Discordant |
| PPP6C | LoF | LoF | Concordant |
| PTEN | LoF | LoF | Concordant |
| PTPN11 | GoF | GoF | Concordant |
| PTPRT | LoF | LoF | Concordant |
| RAC1 | GoF | GoF, LoF | Semi-concordant |
| RAD50 | LoF | LoF | Concordant |
| RAF1 | GoF | GoF | Concordant |
| RARA | GoF | GoF | Concordant |
| RB1 | LoF | LoF | Concordant |
| RBM10 | LoF | LoF | Concordant |
| RET | GoF | GoF | Concordant |
| RIT1 | GoF | GoF | Concordant |
| RNF43 | LoF | LoF | Concordant |
| RPS6KA4 | LoF | LoF | Concordant |
| RUNX1 | GoF, LoF | GoF, LoF | Concordant |
| SDHA | LoF | LoF | Concordant |
| SDHAF2 | LoF | LoF | Concordant |
| SESN2 | GoF, LoF | GoF, LoF | Concordant |
| SETD2 | LoF | LoF | Concordant |
| SMAD2 | LoF | LoF | Concordant |
| SMAD3 | LoF | GoF, LoF | Semi-concordant |
| SMAD4 | LoF | GoF | Discordant |
| SMARCA4 | LoF | GoF | Discordant |
| SMARCB1 | LoF | GoF | Discordant |
| SMARCD1 | LoF | LoF | Concordant |
| SMO | GoF | LoF | Discordant |
| SOCS1 | LoF | LoF | Concordant |
| SOS1 | GoF | GoF | Concordant |
| SOX17 | LoF | LoF | Concordant |
| SPOP | LoF | GoF | Discordant |
| SPRED1 | LoF | LoF | Concordant |
| STAG2 | LoF | LoF | Concordant |
| STAT3 | GoF | GoF | Concordant |
| STK11 | LoF | LoF | Concordant |
| SUZ12 | GoF, LoF | LoF | Semi-concordant |
| TBX3 | GoF, LoF | LoF | Semi-concordant |
| ELOC | LoF | LoF | Concordant |
| TCF3 | GoF, LoF | LoF | Semi-concordant |
| TGFBR1 | GoF | GoF, LoF | Semi-concordant |
| TGFBR2 | LoF | LoF | Concordant |
| TNFRSF14 | LoF | LoF | Concordant |
| TP53 | GoF, LoF | LoF | Semi-concordant |
| TP63 | GoF, LoF | GoF, LoF | Concordant |
| U2AF1 | GoF | GoF | Concordant |
| VHL | LoF | LoF | Concordant |
| XPO1 | GoF | GoF | Concordant |

Supplemental Table 3. Odds ratio scores for classifying germline missense variants in ClinVar and their overlap with cancer mutations in Cancer Hotspots.

| Variant classification/type | Cancer hotspot | Non-cancer hotspot | OR*^a^* | CI*^b^* |
| --- | --- | --- | --- | --- |
| LP/P | 426 | 2723 | — | — |
| LB/B | 4 | 2751 | 107.6^***^ | 40.1-288.4 |
| VUS | 261 | 45181 | — | — |
| LB/B + VUS | 265 | 47932 | 28.3^***^ | 24.2-33.1 |
| LB/B + VUS + CIP | 379 | 50870 | 21.0^***^ | 18.2-24.2 |

*^a^* The three odds ratios (OR) compare likely pathogenic (LP)/pathogenic (P) variants with likely benign (LB)/benign (B), LB/B + variant of uncertain significance (VUS), and LB/B + VUS + conflicting interpretations of pathogenicity (CIP) variants.

*^b^* 95% confidence interval (CI)

^***^ p<0.001

Supplemental Table 4. Positive likelihood ratios for classifying germline missense variants in ClinVar and their overlap with cancer mutations in Cancer Hotspots.

| Variant classification/type | Cancer hotspot | Non-cancer hotspot | LR+*^a^* |
| --- | --- | --- | --- |
| LP/P | 426 | 2723 | — |
| LB/B + VUS | 265 | 47932 | 11.5 |

*^a^* Positive likelihood ratio (LR+) compare likely pathogenic (LP)/pathogenic (P) variants with likely benign (LB)/benign (B) and LB/B + variant of uncertain significance (VUS) variants.

**Supplemental Table 5.** Variants from controlled-access databases that overlapped with cancer mutations from Cancer Hotspots, along with participant counts and ClinVar classifications.

|  | Total participants sequenced*^a^* | Total participants with matches^b^ | Total variant  matches | Variant classification in ClinVar*^c^* | | | |
| --- | --- | --- | --- | --- | --- | --- | --- |
|  |  |  |  | LP/P | LB/B | VUS | N/P |
| GEL | 1,048,576 | 334 | 144 | 46 | 1 | 29 | 68 |
| MSSNG | 11,312 | 21 | 15 | 3 | 0 | 8 | 4 |
| G4RD | 2,799 | 24 | 17 | 3 | 0 | 2 | 12 |
| GeneDx | 400,000 | 1,296 | 175 | 111 | 0 | 27 | 37 |
| Total | 1,462,687 | 1,675 | 351 | 163 | 1 | 66 | 121 |
| **Unique** | **1,462,687** | **1,675** | **302** | **140** | **1** | **53** | **108** |

*^a^*Total number of participants in the queried database.

*^b^*Participants with variants overlapping cancer mutations from Cancer Hotspots.

*^c^*Overlapping variants with ClinVar information: LP/P, LB/B, VUS, N/P (absent in ClinVar).

**Supplemental Figure 1.** Workflow for extracting germline missense variants from ClinVar found in the list of 216 genes from Cancer Hotspots. This illustrates the process of filtering the variants to create the “ClinVar dataset” used in the odd ratio calculations and as the training dataset for supervised learning models.

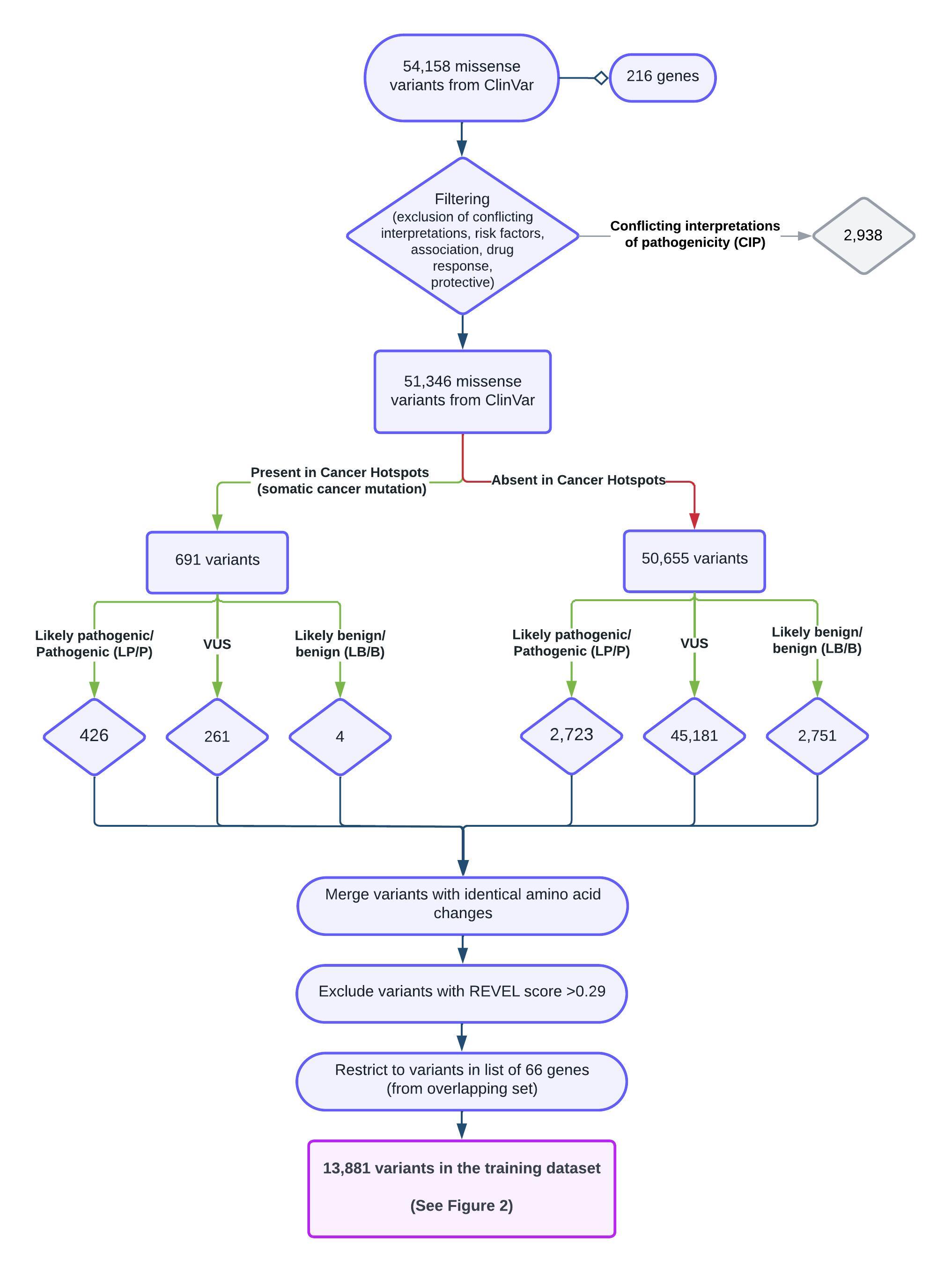

**Supplemental Figure 2.** Bar graph showing the distribution of cancer gene types, categorized as proto-oncogenes, tumor suppressor genes (TSGs), dual-function (both proto-oncogene and TSG) or not yet defined by the Cancer Gene Census. (A) Distribution for the 216 cancer genes included in the Cancer Hotspots database. (B) Distribution for 84 cancer genes with mutations that overlap with germline variants in ClinVar.

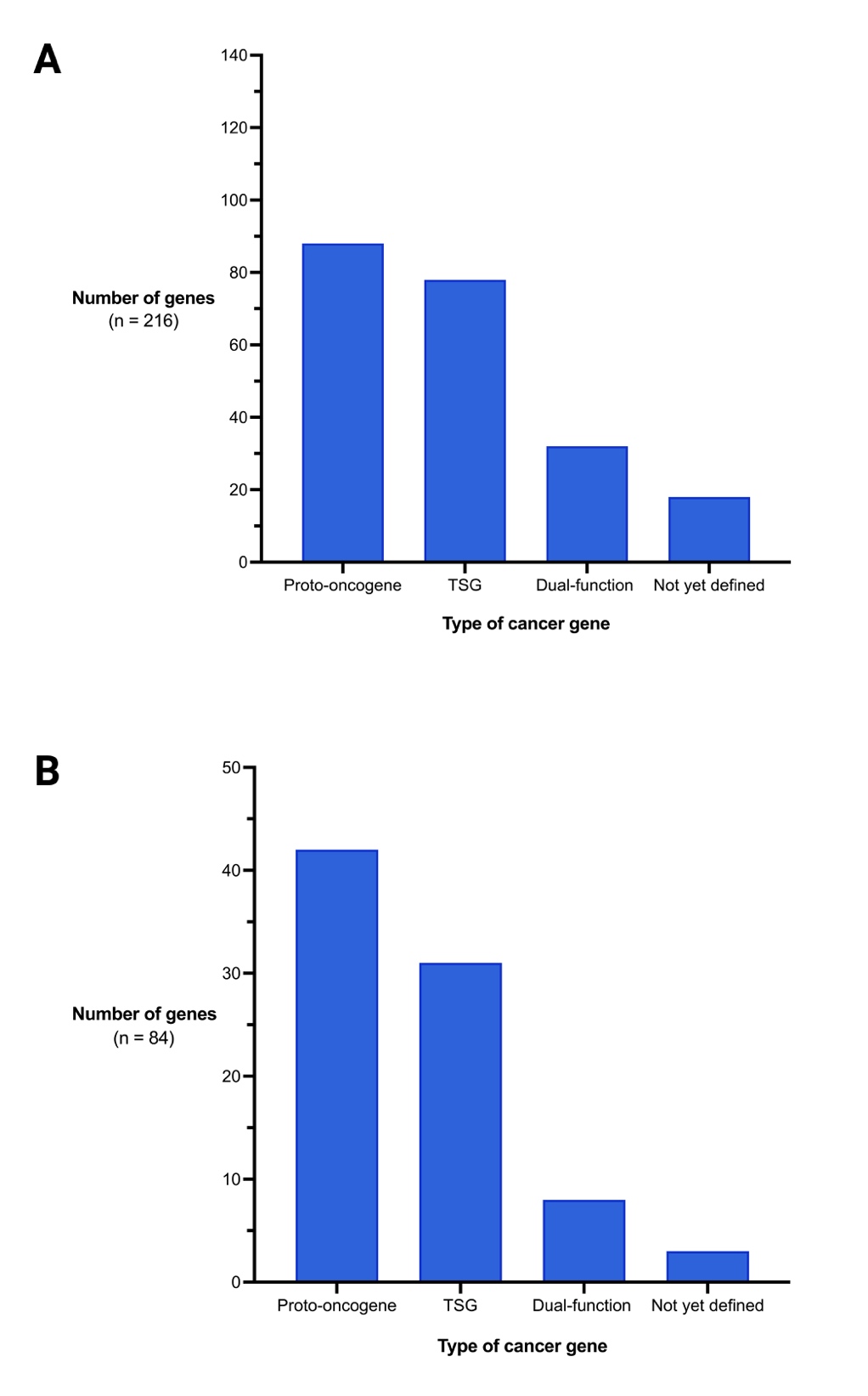

**Supplemental Figure 3.** Functional impact of missense cancer mutations from Cancer Hotspots determined using the Clinical Knowledgebase (CKB)^1^ and germline variant classifications in ClinVar. GoF, gain-of-function; LoF loss-of-function.

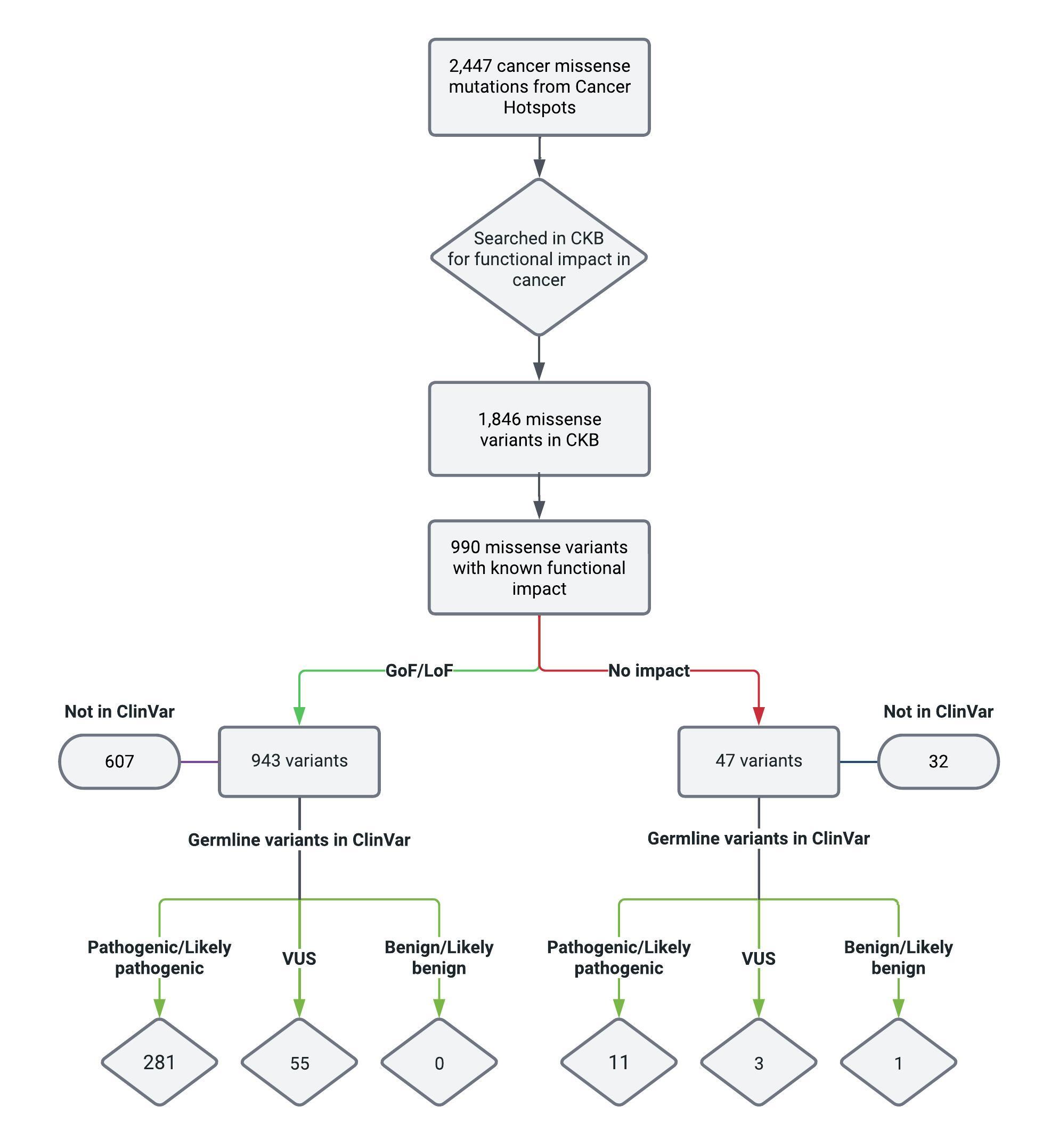

**Supplemental Figure 4.** Hypothetical impact of applying an additional pathogenic moderate (PM) evidence-level criterion to the interpretation of germline variants of uncertain significance (VUS) in ClinVar that overlap with cancer mutations from Cancer Hotspots. Each row represents the existing evidence codes for VUS, with the addition of one PM criterion, to form a combining criterion for the classification of “likely pathogenic” according to the ACMG/AMP guidelines.^7^ Among the 261 VUS, 12 were recently reclassified to LP/P (n=11) or LB (n =1) in ClinVar. With the remaining 249, an additional PM evidence code would be enough to potentially upgrade 66 VUS (26.5%) to LP. Figure was created with BioRender and adapted from Brnich et al., (2018).^2^

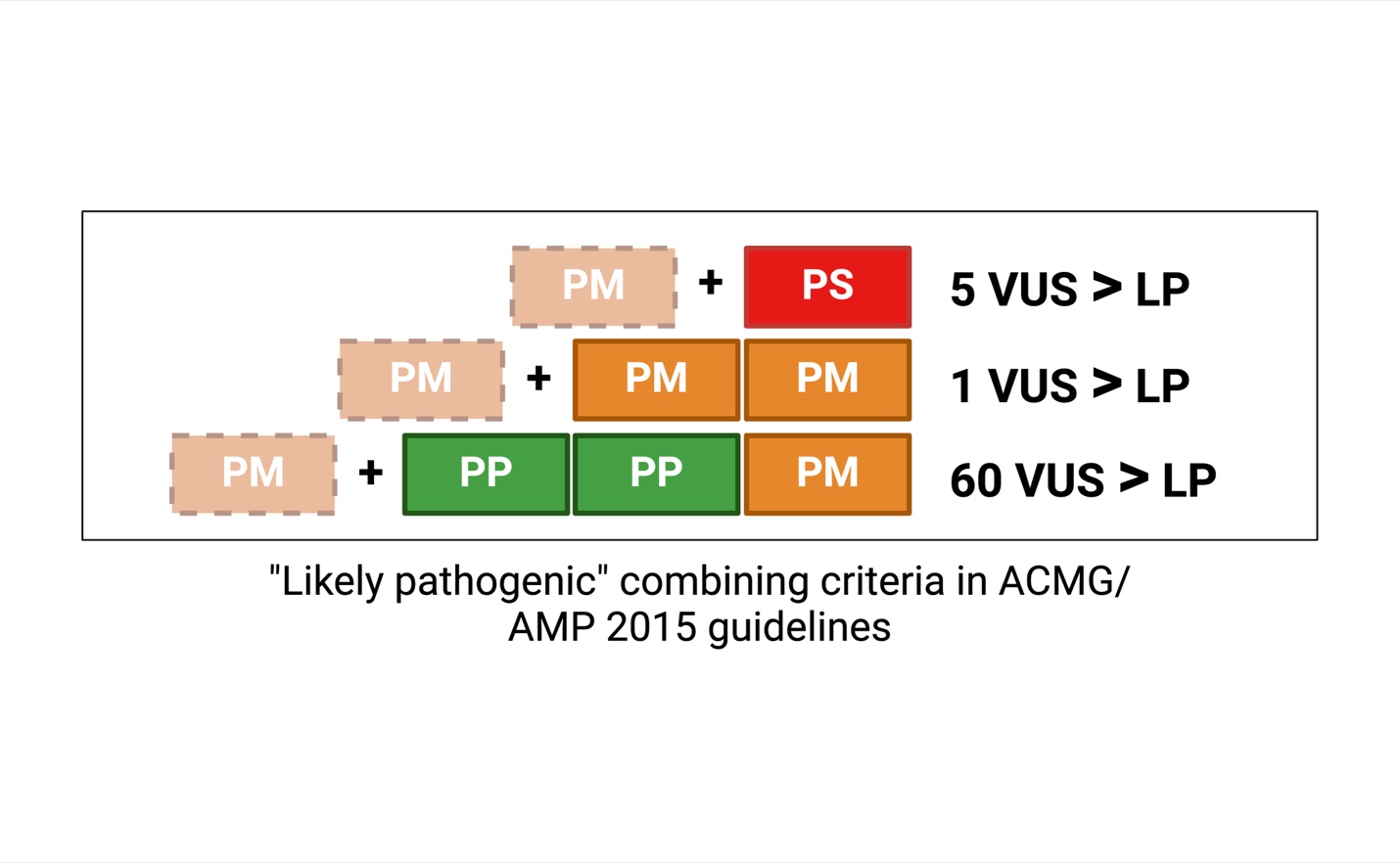

**Supplemental Figure 5.** Distribution of (A) REVEL^4^ and (B) AlphaMissense^5^ scores for CH cancer mutations (n = 2,447) and variants in the ClinVar dataset (n = 51,346). Variants are categorized by the presence of an overlap with cancer mutations from Cancer Hotspots and absence from Cancer Hotspots. The figure includes a category for cancer mutations from Cancer Hotspots not reported in ClinVar (ClinVar absent). The median scores for each group are indicated on the plot. Score thresholds corresponding to various PP3 and BP4 evidence strengths are displayed by labels above the dotted lines.

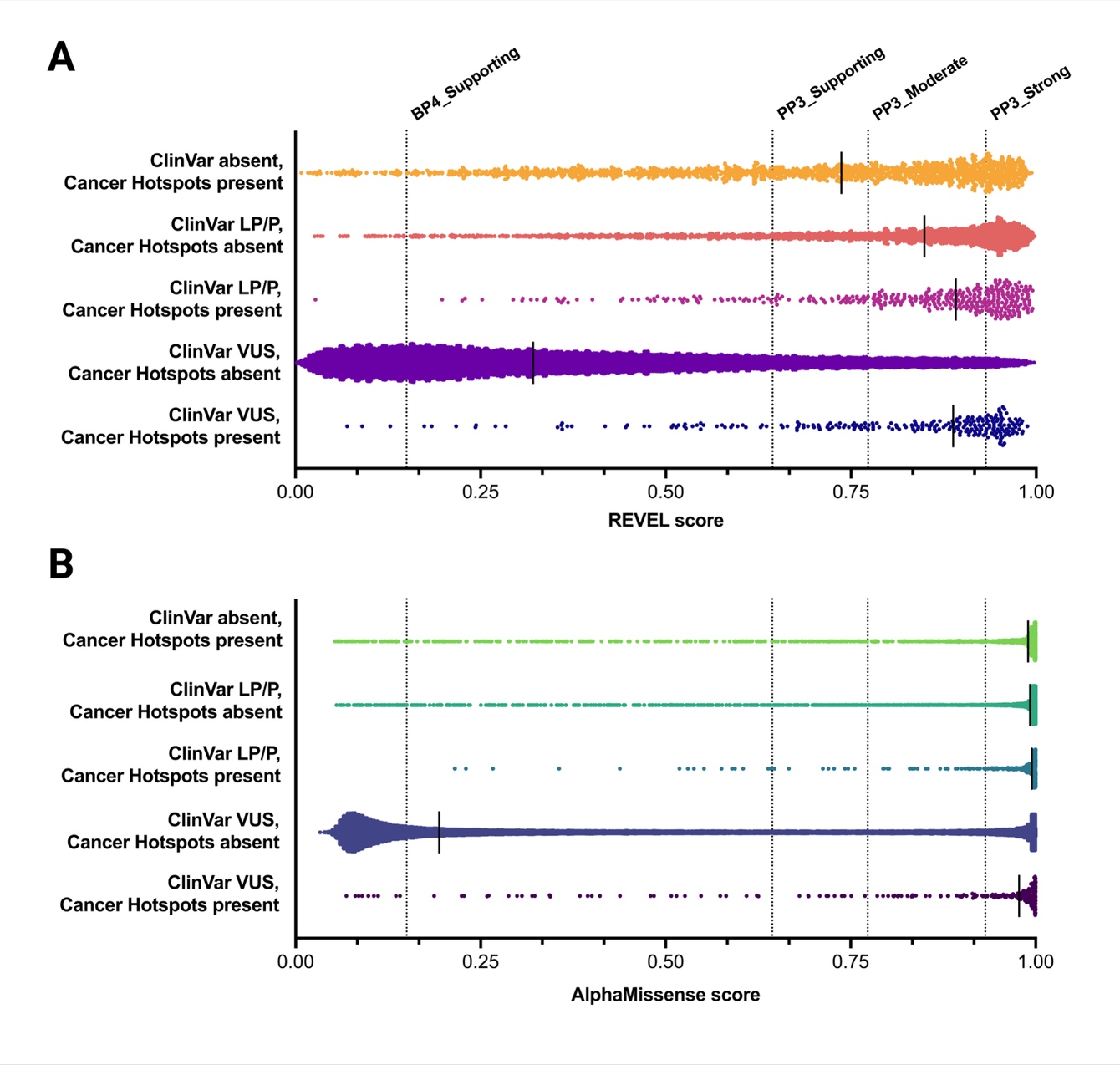

**Supplemental Figure 6.** Workflow for obtaining confirmed somatic missense mutations from the COSMIC Cancer Gene Census coding mutations. This figure illustrates the process of filtering COSMIC mutations using a stringent tumor sample count filter to identify additional cancer mutations that are absent from Cancer Hotspots.

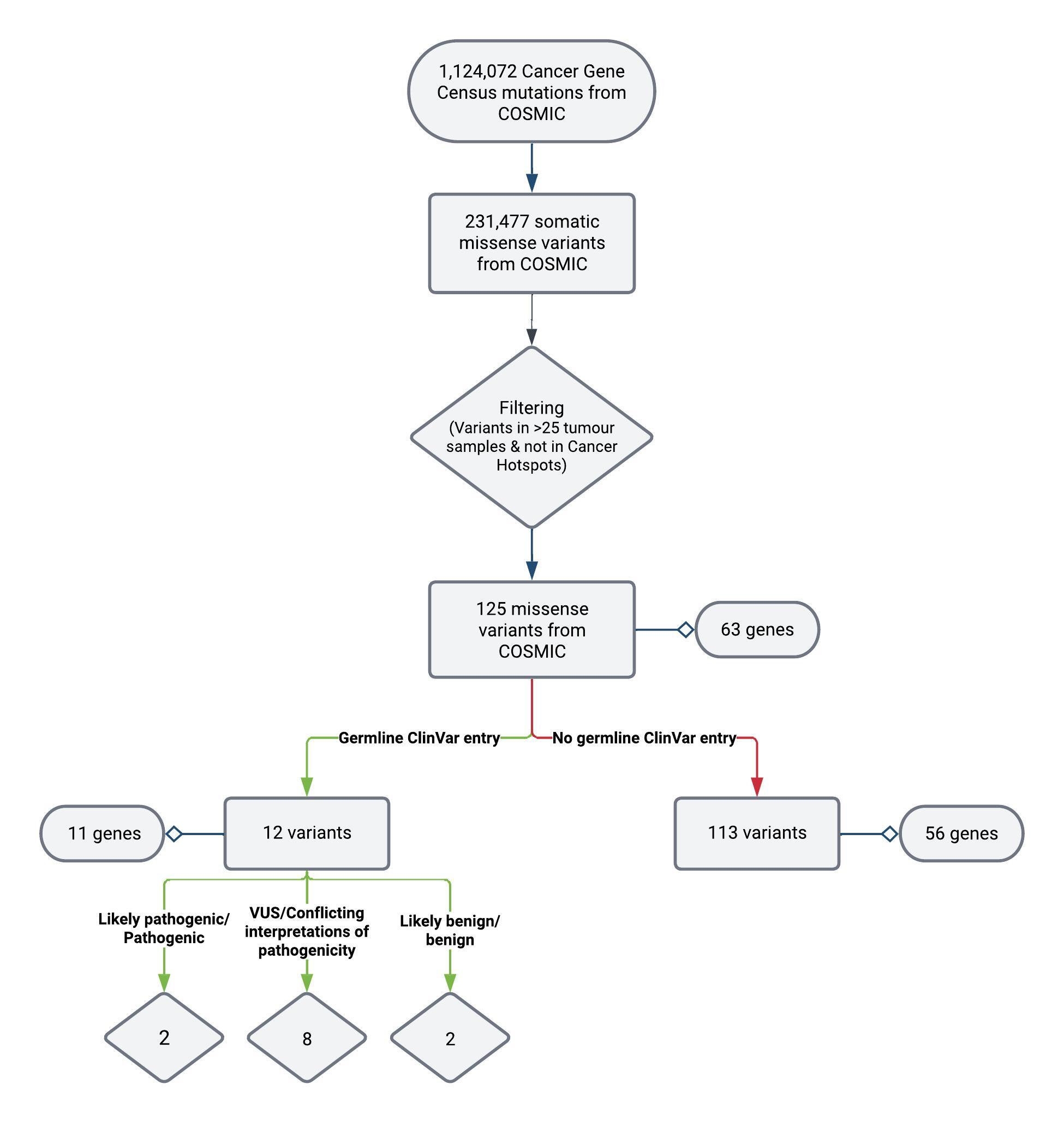

**Supplemental Figure 7.** Proportion of cancer missense mutations from Cancer Hotspots reported in ClinVar for a subset of genes for *TP53*, *PIK3CA*, *PTEN*, *SMAD4*, *VHL*, *PTPN11*, *RIT1*, and *FGFR3.* Mutations that are present in ClinVar are indicated in blue, absent are indicated in purple. LP/P variants (pink) and LB/B/VUS (orange) variants among those present in ClinVar are also shown.

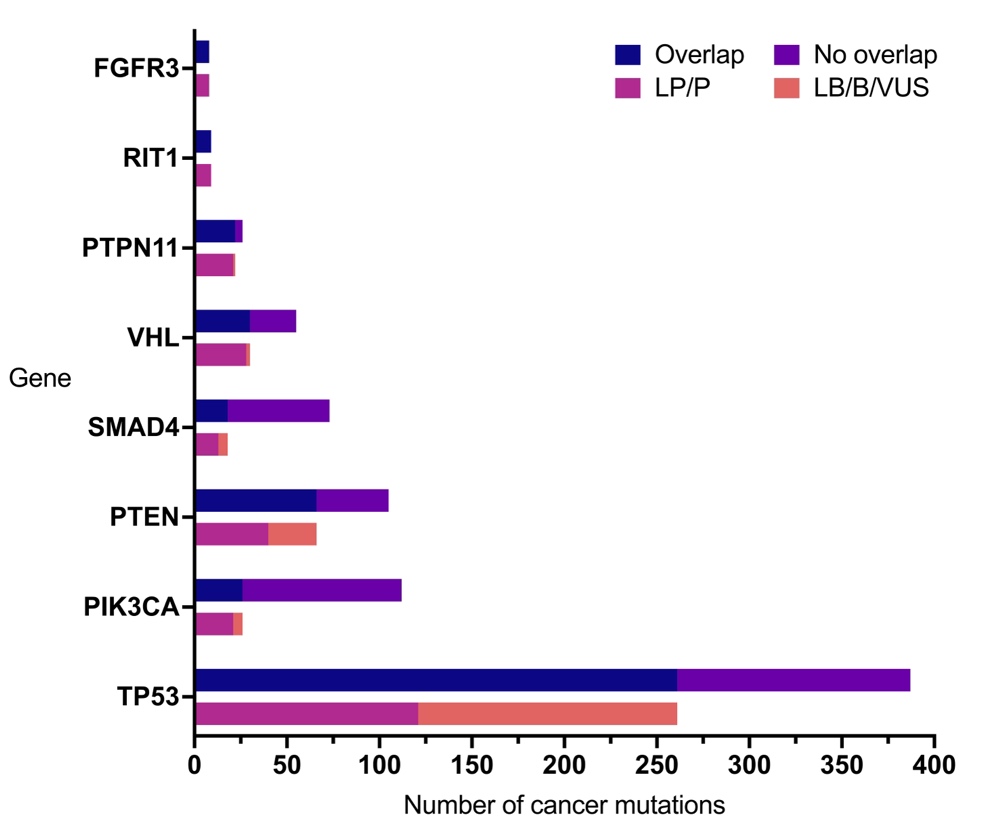

**Supplemental Figure 8.** Number of tumor samples with cancer mutations from Cancer Hotspots and their germline variant classifications. (A) Tumor sample count for LP/P variants compared to LB/B/VUS variants, with an ROC curve that evaluates the discriminatory power between the two groups based on tumor sample counts. LP/P variants exhibited significantly higher tumor sample counts than LB/B/VUS variants (p < 0.0001). The ROC curve yielded an area under the curve (AUC) value of 0.614, indicating moderate discriminatory ability. (B) Tumor sample counts for the same analysis as (A) but restricted to sample counts >25. his subset analysis revealed higher discriminatory median counts (54 and 32.5 samples for LP/P and LB/B/VUS groups, respectively) and achieved the largest AUC of 0.7640. Mann-Whitney U test was performed to assess the statistical difference between LP/P and LB/B/VUS.

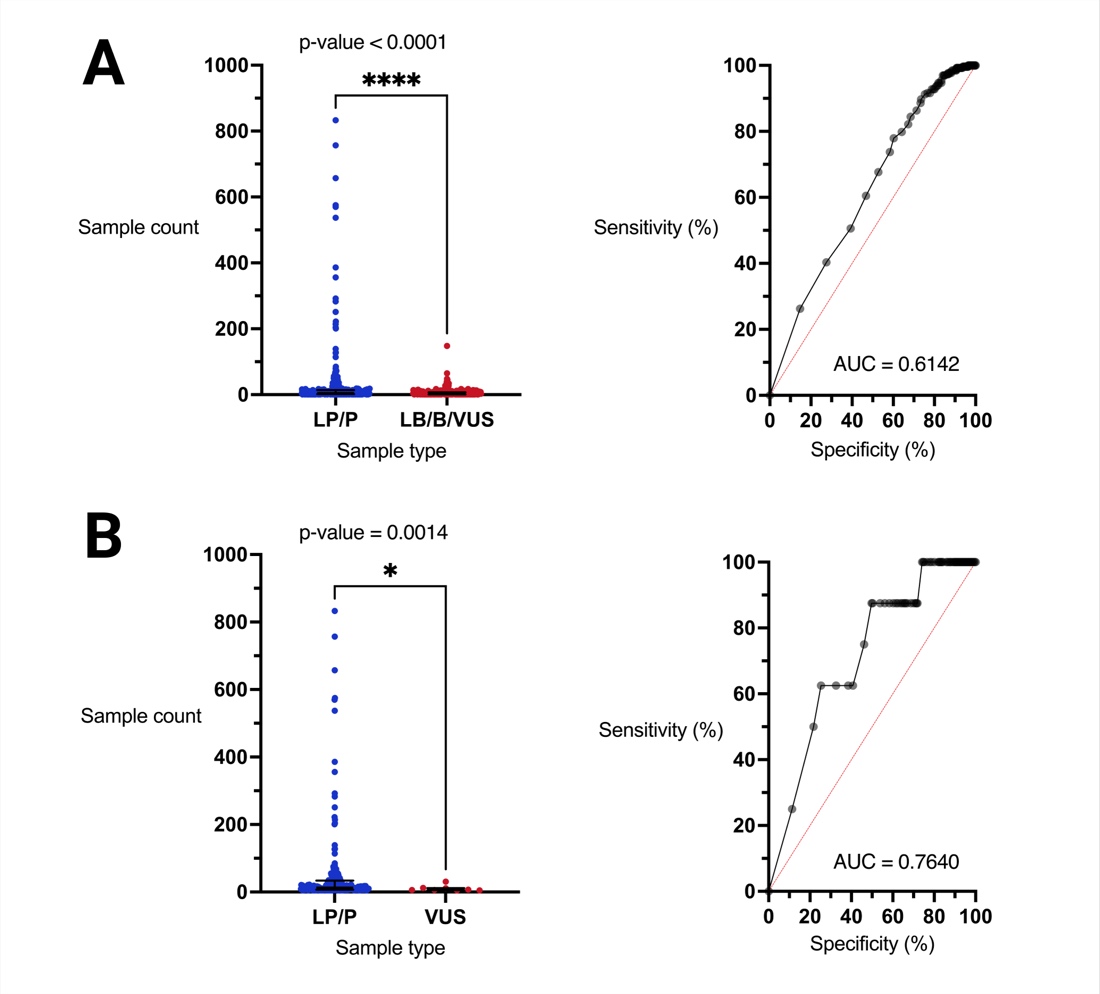

**Supplemental Figure 9.** Distribution of conservation scores for germline missense variants annotated with (A) phastCons and (B) phyloP across vertebrate and mammalian species. Variants are categorized by the presence of an overlap with cancer mutations from Cancer Hotspots and absence from Cancer Hotspots. The figure includes a category for cancer mutations from Cancer Hotspots not reported in ClinVar (ClinVar absent). The median scores for each group are indicated on the plot. Mann-Whitney U test was performed to assess the differences between Cancer Hotspots absent and present variants, and the probability of superiority (PS) was calculated to determine effect size.

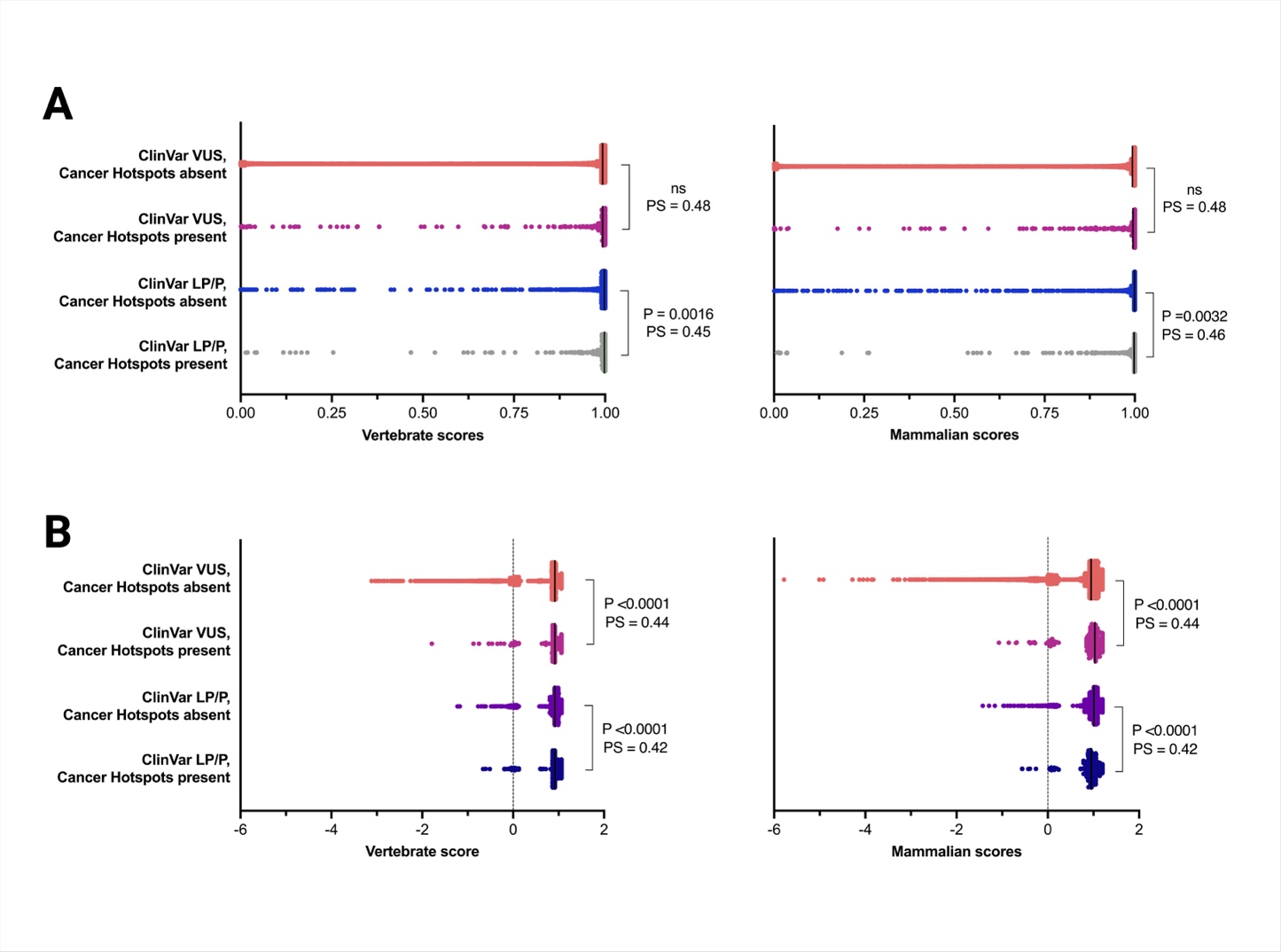

**Supplemental Figure 10.** Receiver-operating characteristic (ROC) curve comparing the performance of the logistic regression model (blue) and the random forest model (purple) using the (A) test dataset and (B) cross-validation set. The models' performance was evaluated using k-fold cross-validation, with k=8 for logistic regression and k=10 for random forest. AUC, area under the curve.

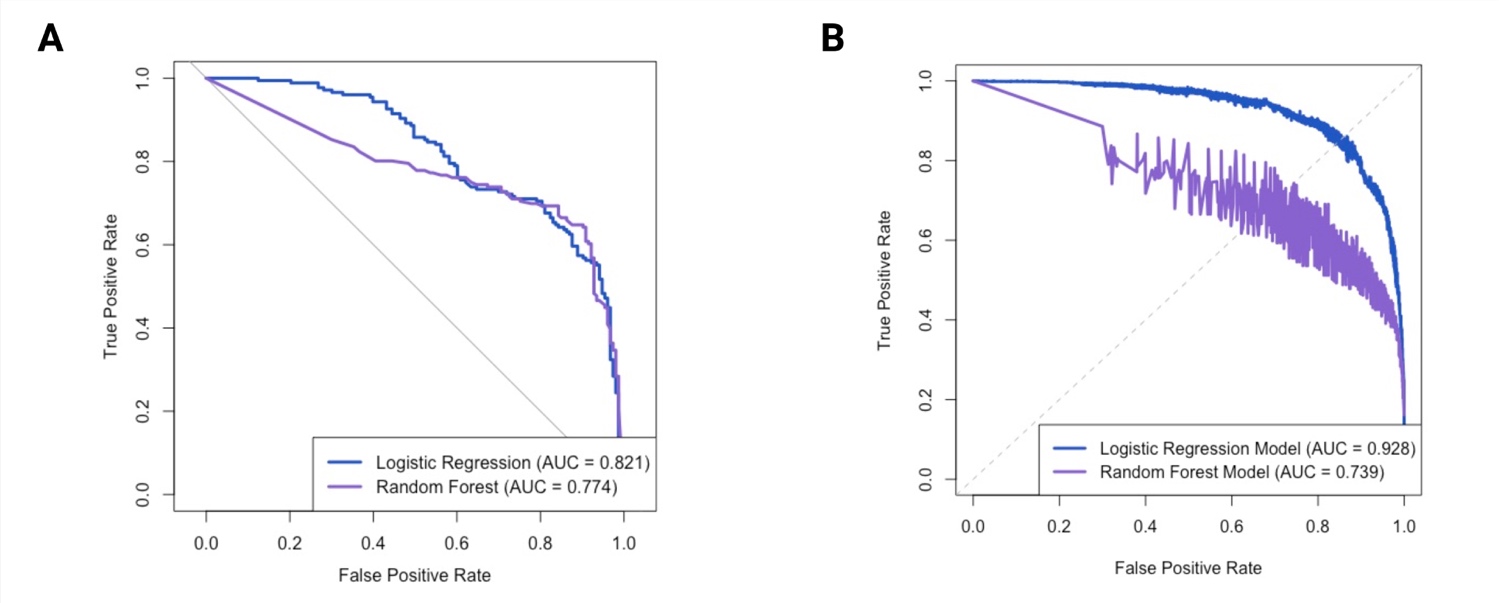

**Supplemental Figure 11.** Accuracy of supervised learning models with test dataset using optimal thresholds. (A) Confusion matrix showing the correctly classified variants (green) and incorrectly classified variants (red) by the logistic regression model using an optimal threshold of 0.74. (B) Confusion matrix showing variant classification by the random forest model using an optimal threshold of 0.39. AUC, area under the curve; LB/B, Likely benign/Benign; LP/P, Likely pathogenic/Pathogenic; VUS, variant of uncertain significance. Created with R and BioRender.

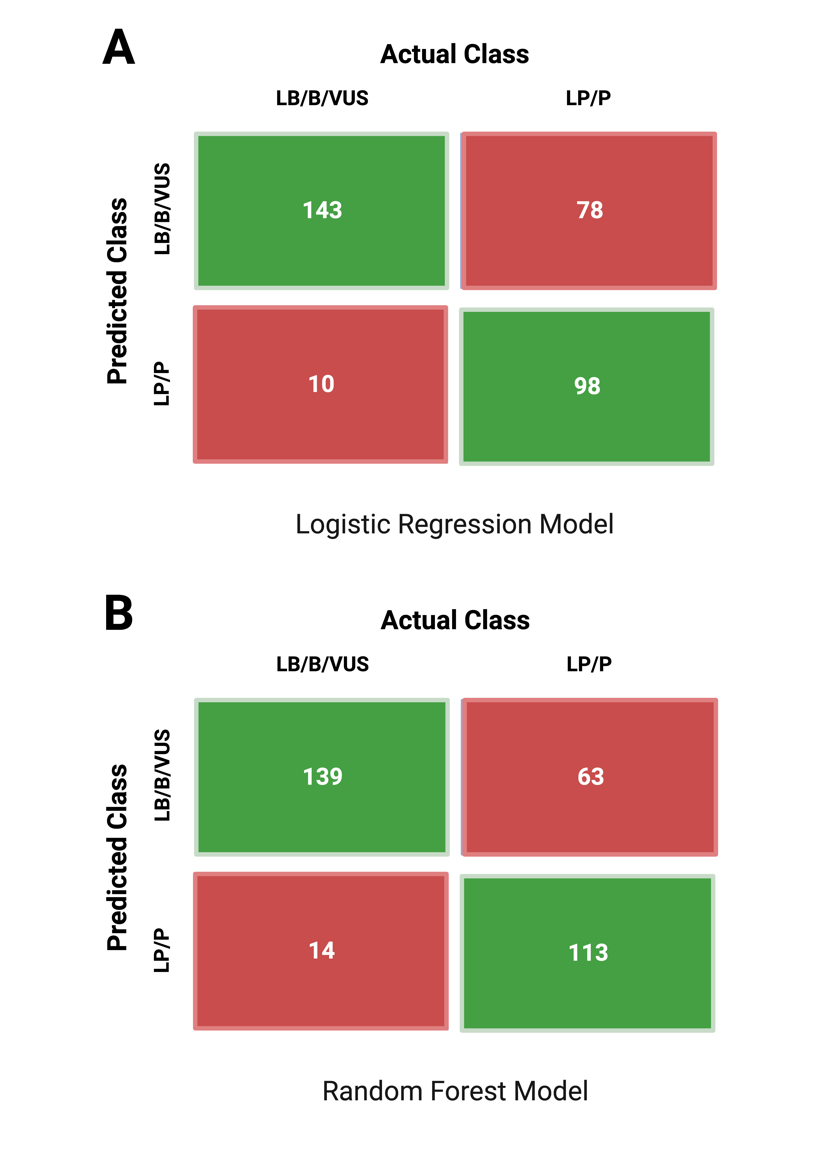

**Supplemental Figure 12.** Comparisons of pathogenicity scores for LRM, RFM, and other known *in silico* prediction tools and performance on predicting pathogenicity on test dataset (n=339). (A) Bar graph showing the area under the precision-recall curve (AUPRC) for each tool, including logistic regression model (LRM), random forest model (RFM), SIFT, PolyPhen-2, REVEL, CADD, VARITY, AlphaMissense, PrimateAI, VEST4, and MutPred2. (B) Precision-recall curves for first-generation tools (SIFT and PolyPhen-2) compared with LRM and RFM. (C) Precision-recall curves for second-generation tools (REVEL, CADD, VARITY, VEST4) compared with LRM and RFM. (D) Precision-recall curves for third-generation tools (AlphaMissense,PrimateAI, and MutPred2) compared with LRM and RFM.

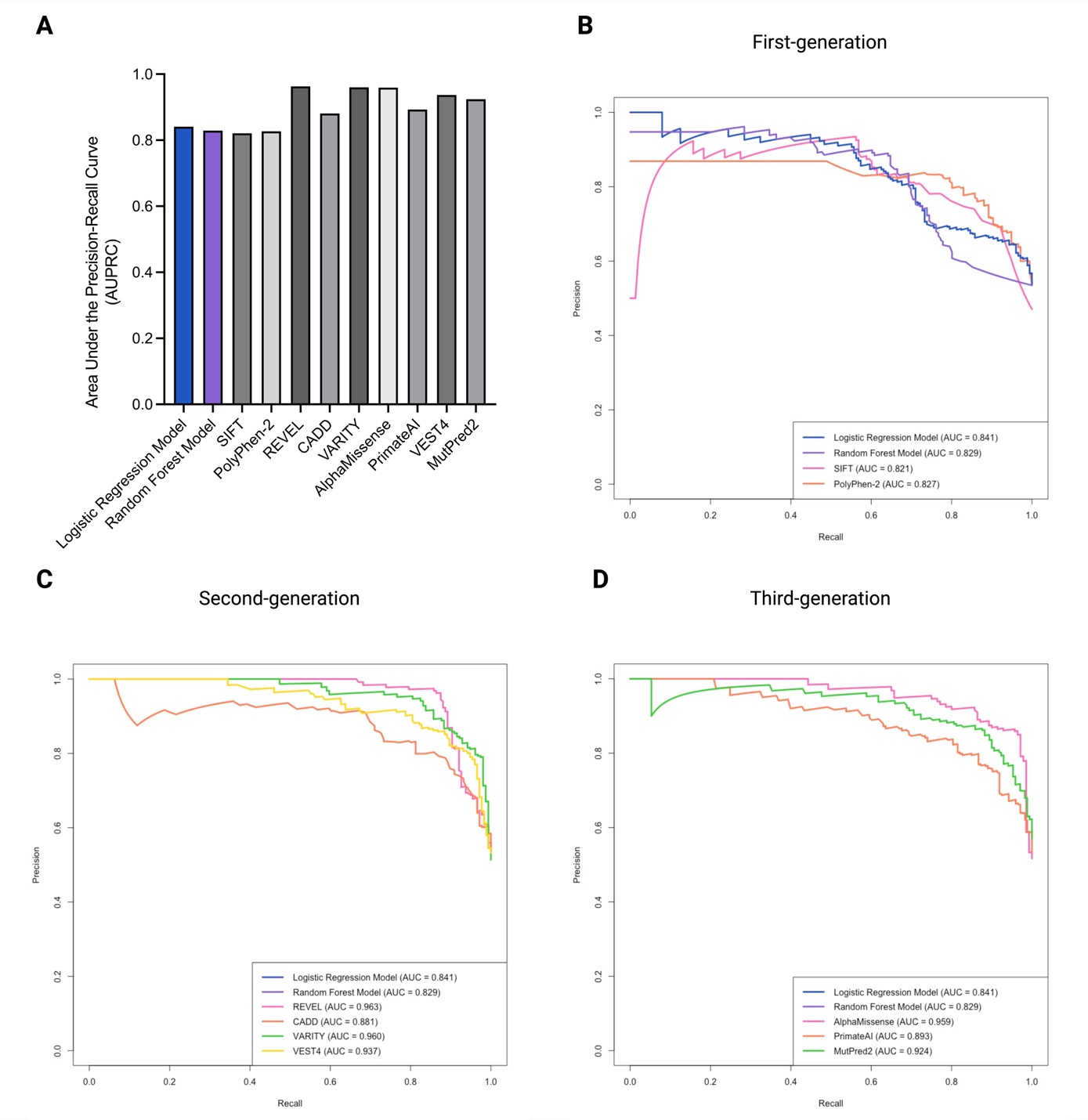

**Supplemental Appendix 1.** R script used to train and test logistic regression model.

```
# Load necessary libraries
library(ggplot2)
library(PRROC)

# Load and preprocess the training data
df <- read.csv("your_file_name.csv", header = TRUE)

# Filter data to exclude rows where REVEL > 0.290 for VUS classification
df <- df[!(df$clinvar_class == "VUS" & df$REVEL > 0.290), ]

# Generate a list of genes associated with cancer
genes_list <- unique(df$gene[df$cancer_binary == "1"])

# Subset df to include only genes in genes_list
df <- df[df$gene %in% genes_list, ]

# Convert specific columns to factors
factor_columns <- c("gene", "gene_residue", "residue", "cancer_binary", "outcome_binary", "clinvar_class", "allele_categorical")
df[factor_columns] <- lapply(df[factor_columns], as.factor)

# Remove rows with missing values in key predictive columns
df <- df[complete.cases(df[, c("phyloP7way_vertebrate", "phastCons7way_vertebrate", "REVEL")]), ]

# Build logistic regression model to predict binary outcome
model <- glm(outcome_binary ~ gene + residue_count + cancer_binary + cancer_count +
             phyloP7way_vertebrate + phyloP20way_mammalian +
             phastCons7way_vertebrate + phastCons20way_mammalian,
             family = "binomial", data = df, weights = df$wgts)

# Summarize model results without scientific notation
options(scipen = 999)
summary(model)

# Load and preprocess the test dataset
dfTest <- read.csv("your_file_name.csv")

# Function to convert data types for specified columns
convertDataType <- function(df, columns, type) {
  df[columns] <- lapply(df[columns], type)
  return(df)
}

# Convert categorical columns to factors and numerical columns to numeric
dfTest <- convertDataType(dfTest,
                          c("gene", "gene_residue", "residue", "cancer_binary", "clinvar_class"),
                          as.factor)
dfTest <- convertDataType(dfTest,
                          c("phyloP7way_vertebrate", "phyloP20way_mammalian",
                            "phastCons7way_vertebrate", "phastCons20way_mammalian",
                            "REVEL", "SIFT", "PolyPhen2", "AlphaMissense"),
                          as.numeric)

# Replace "NA" strings with actual NA values and remove incomplete rows
dfTest[dfTest == "NA"] <- NA
dfTest <- dfTest[complete.cases(dfTest[c("phyloP7way_vertebrate", "phastCons7way_vertebrate")]), ]

# Predict outcome probabilities on the test dataset
test_pred_prob <- predict(model, newdata = dfTest, type = "response")

# Separate predicted probabilities by class (positive: outcome_binary=1, negative: outcome_binary=0)
scores.class0 <- test_pred_prob[dfTest$outcome_binary == "1"]
scores.class1 <- test_pred_prob[dfTest$outcome_binary == "0"]

# Plot Precision-Recall curve
prc <- pr.curve(scores.class0, scores.class1, curve = TRUE)
plot(prc, ylim = c(0, 1), xlim = c(0, 1), col = "#1F57C3", lwd = 3,
```

main = "Precision-Recall Curve", xlab = "Recall", ylab = "Precision")

**Supplemental Appendix 2.** R script used to train and test random forest model.

```
# Load necessary libraries
library(randomForest)
library(PRROC)

# Load and preprocess the training dataset
df <- read.csv("your_file_name.csv", header = TRUE)

# Filter data to exclude rows where REVEL > 0.290 for VUS classification
df <- df[!(df$clinvar_class == "VUS" & df$REVEL > 0.290), ]

# Generate a list of genes associated with cancer
genes_list <- unique(df$gene[df$cancer_binary == "1"])

# Subset df to include only genes in genes_list
df <- df[df$gene %in% genes_list, ]

# Convert specific columns to factors
df$cancer_binary <- as.factor(df$cancer_binary)
df$outcome_binary <- as.factor(df$outcome_binary)

# Remove rows with missing values in key predictive columns
df <- df[complete.cases(df[, c("phyloP7way_vertebrate", "phastCons7way_vertebrate")]), ]

# Build random forest model for binary outcome prediction
set.seed(42) # Ensures reproducibility
rf.model <- randomForest(outcome_binary ~ residue_count + cancer_binary + cancer_count +
                         phyloP7way_vertebrate + phyloP20way_mammalian +
                         phastCons7way_vertebrate + phastCons20way_mammalian,
                         data = df, mtry = 4, importance = TRUE, ntree = 350, proximity = FALSE)

# Load and preprocess the test dataset
dfTest <- read.csv("your_file_name.csv", header = TRUE)

# Convert relevant columns to factors in test data
factor_columns_test <- c("gene", "gene_residue", "residue", "cancer_binary", "clinvar_class", "outcome_binary")
dfTest[factor_columns_test] <- lapply(dfTest[factor_columns_test], as.factor)

# Remove rows with missing values in relevant columns and subset by genes_list
dfTest <- dfTest[complete.cases(dfTest[, c("phyloP7way_vertebrate", "phastCons7way_vertebrate", "REVEL")]), ]
dfTest <- dfTest[dfTest$gene %in% genes_list, ]

# Predict outcome probabilities on the test dataset using random forest model
test_pred_prob_rf <- predict(rf.model, dfTest, type = "prob")[, 2]

# Separate predicted probabilities by class (positive: outcome_binary=1, negative: outcome_binary=0)
scores.class0_rf <- test_pred_prob_rf[dfTest$outcome_binary == "1"]
scores.class1_rf <- test_pred_prob_rf[dfTest$outcome_binary == "0"]

# Plot Precision-Recall curve for random forest model
prc_rf <- pr.curve(scores.class0_rf, scores.class1_rf, curve = TRUE)
plot(prc_rf, ylim = c(0, 1), xlim = c(0, 1), col = "#8862CE", lwd = 3,
     main = "Precision-Recall Curve", xlab = "Recall", ylab = "Precision")
```

lines(prc_rf$curve[,1], prc_rf$curve[,2], col = "#8862CE", lwd = 3)

**List of Legends**

Supplemental Table 1. Genes with missense mutations from Cancer Hotspots that overlap with germline variants in ClinVar, along with their OMIM disease phenotype(s).

**Supplemental Table 2.** Genes from Cancer Hotspots with known modes of inheritance for associated Mendelian disease(s), including somatic and germline mechanisms of action and concordance status (concordant, discordant, or semi-concordant).

**Supplemental Table 3.** Odds ratio scores for classifying germline missense variants in ClinVar and their overlap with cancer mutations in Cancer Hotspots.

**Supplemental Table 4.** Positive likelihood ratios for classifying germline missense variants in ClinVar and their overlap with cancer mutations in Cancer Hotspots.

**Supplemental Table 5.** Variants from controlled-access databases that overlapped with cancer mutations from Cancer Hotspots, along with participant counts and ClinVar classifications.

**Supplemental Figure 1.** Workflow for extracting germline missense variants from ClinVar found in the list of 216 genes from Cancer Hotspots. This illustrates the process of filtering the variants to create the “ClinVar dataset” used in the odd ratio calculations and as the training dataset for supervised learning models.

**Supplemental Figure 2.** Bar graph showing the distribution of cancer gene types, categorized as proto-oncogenes, tumor suppressor genes (TSGs), dual-function (both proto-oncogene and TSG) or not yet defined by the Cancer Gene Census. (A) Distribution for the 216 cancer genes included in the Cancer Hotspots database. (B) Distribution for 84 cancer genes with mutations that overlap with germline variants in ClinVar.

**Supplemental Figure 3.** Functional impact of missense cancer mutations from Cancer Hotspots determined using the Clinical Knowledgebase (CKB)^1^ and germline variant classifications in ClinVar.  GoF, gain-of-function; LoF loss-of-function.

**Supplemental Figure 4.** Hypothetical impact of applying an additional pathogenic moderate (PM) evidence-level criterion to the interpretation of germline variants of uncertain significance (VUS) in ClinVar that overlap with cancer mutations from Cancer Hotspots. Each row represents the existing evidence codes for VUS, with the addition of one PM criterion, to form a combining criterion for the classification of “likely pathogenic” according to the ACMG/AMP guidelines.^7^ Among the 261 VUS, 12 were recently reclassified to LP/P (n=11) or LB (n =1) in ClinVar. With the remaining 249, an additional PM evidence code would be enough to potentially upgrade 66 VUS (26.5%) to LP. Figure was created with BioRender and adapted from Brnich et al., (2018).^2^

**Supplemental Figure 5.** Distribution of (A) REVEL^4^ and (B) AlphaMissense^5^ scores for CH cancer mutations (n = 2,447) and variants in the ClinVar dataset (n = 51,346). Variants are categorized by the presence of an overlap with cancer mutations from Cancer Hotspots and absence from Cancer Hotspots. The figure includes a category for cancer mutations from Cancer Hotspots not reported in ClinVar (ClinVar absent). The median scores for each group are indicated on the plot. Score thresholds corresponding to various PP3 and BP4 evidence strengths are displayed by labels above the dotted lines.

**Supplemental Figure 6.** Workflow for obtaining confirmed somatic missense mutations from the COSMIC Cancer Gene Census coding mutations. This figure illustrates the process of filtering COSMIC mutations using a stringent tumor sample count filter to identify additional cancer mutations that are absent from Cancer Hotspots.

**Supplemental Figure 7.** Proportion of cancer missense mutations from Cancer Hotspots reported in ClinVar for a subset of genes for *TP53*, *PIK3CA*, *PTEN*, *SMAD4*, *VHL*, *PTPN11*, *RIT1*, and *FGFR3.* Mutations that are present in ClinVar are indicated in blue, absent are indicated in purple. LP/P variants (pink) and LB/B/VUS (orange) variants among those present in ClinVar are also shown.

**Supplemental Figure 8.** Number of tumor samples with cancer mutations from Cancer Hotspots and their germline variant classifications. (A) Tumor sample count for LP/P variants compared to LB/B/VUS variants, with an ROC curve that evaluates the discriminatory power between the two groups based on tumor sample counts. LP/P variants exhibited significantly higher tumor sample counts than LB/B/VUS variants (p < 0.0001). The ROC curve yielded an area under the curve (AUC) value of 0.614, indicating moderate discriminatory ability. (B) Tumor sample counts for the same analysis as (A) but restricted to sample counts >25. his subset analysis revealed higher discriminatory median counts (54 and 32.5 samples for LP/P and LB/B/VUS groups, respectively) and achieved the largest AUC of 0.7640. Mann-Whitney U test was performed to assess the statistical difference between LP/P and LB/B/VUS.

**Supplemental Figure 9.** Distribution of conservation scores for germline missense variants annotated with (A) phastCons and (B) phyloP across vertebrate and mammalian species. Variants are categorized by the presence of an overlap with cancer mutations from Cancer Hotspots and absence from Cancer Hotspots. The figure includes a category for cancer mutations from Cancer Hotspots not reported in ClinVar (ClinVar absent). The median scores for each group are indicated on the plot. Mann-Whitney U test was performed to assess the differences between Cancer Hotspots absent and present variants, and the probability of superiority (PS) was calculated to determine effect size.

**Supplemental Figure 10.** Receiver-operating characteristic (ROC) curve comparing the performance of the logistic regression model (blue) and the random forest model (purple) using the (A) test dataset and (B) cross-validation set. The models' performance was evaluated using k-fold cross-validation, with k=8 for logistic regression and k=10 for random forest. AUC, area under the curve.

**Supplemental Figure 11.** Accuracy of supervised learning models with test dataset using optimal thresholds. (A) Confusion matrix showing the correctly classified variants (green) and incorrectly classified variants (red) by the logistic regression model using an optimal threshold of 0.74. (B) Confusion matrix showing variant classification by the random forest model using an optimal threshold of 0.39. AUC, area under the curve; LB/B, Likely benign/Benign; LP/P, Likely pathogenic/Pathogenic; VUS, variant of uncertain significance. Created with R and BioRender.

**Supplemental Figure 12.** Comparisons of pathogenicity scores for LRM, RFM, and other known *in silico* prediction tools and performance on predicting pathogenicity on test dataset (n=339). (A) Bar graph showing the area under the precision-recall curve (AUPRC) for each tool, including logistic regression model (LRM), random forest model (RFM), SIFT, PolyPhen-2, REVEL, CADD, VARITY, AlphaMissense, PrimateAI, VEST4, and MutPred2. (B) Precision-recall curves for first-generation tools (SIFT and PolyPhen-2) compared with LRM and RFM. (C) Precision-recall curves for second-generation tools (REVEL, CADD, VARITY, VEST4) compared with LRM and RFM. (D) Precision-recall curves for third-generation tools (AlphaMissense,PrimateAI, and MutPred2) compared with LRM and RFM.
